# Patient-specific alterations in blood plasma cfRNA profiles enable accurate classification of cancer patients and controls

**DOI:** 10.1101/2023.05.24.23290388

**Authors:** Annelien Morlion, Philippe Decruyenaere, Kathleen Schoofs, Jasper Anckaert, Justine Nuytens, Eveline Vanden Eynde, Kimberly Verniers, Celine Everaert, Fritz Offner, Jo Van Dorpe, Olivier Thas, Jo Vandesompele, Pieter Mestdagh

## Abstract

Circulating nucleic acids in blood plasma form an attractive resource to study human health and disease. Here, we applied mRNA capture sequencing of blood plasma cell-free RNA from 266 cancer patients and cancer-free controls (discovery n = 208, 25 cancer types; replication n = 58, 3 types). We observed cancer type-specific as well as pan-cancer alterations in cell-free transcriptomes compared to controls. Differentially abundant RNAs were heterogenous among patients and among cohorts, hampering identification of robust cancer biomarkers. Therefore, we developed a novel method that compares each individual cancer patient to a reference control population to identify so-called biomarker tail genes. The number of biomarker tail genes in a sample enables distinguishing an individual cancer patient from controls. The potential of this novel approach was confirmed in additional cohorts of 65 plasma donors (2 lymphoma types) and 24 urine donors (bladder cancer). Together, our findings demonstrate heterogeneity in cell-free RNA alterations among cancer patients and propose that case-specific alterations can be exploited for classification purposes.

## Introduction

Cancer is a complex disease with considerable heterogeneity, both between as well as within cancer types. Finding robust and easily accessible biomarkers is crucial to improve patient well-being and outcome. Such biomarkers can either assist in early detection of cancer, enabling treatment to start at an earlier stage, or they can support diagnosis, treatment selection or response monitoring. As an alternative for biomarkers that require invasive tissue biopsies, liquid biopsy-based biomarkers such as circulating nucleic acids have emerged as a promising tool for cancer diagnosis and monitoring.

To date, most studies on circulating nucleic acids in cancer patients have focused on circulating tumor DNA (ctDNA). The amount of tumor DNA in circulation is highly dependent on the tumor type and burden^1^, suggesting that ctDNA can be less informative for certain cancer types or early-stage cancers. Cell-free RNA (cfRNA) may complement ctDNA by reflecting dynamic changes in gene expression during cancer development and progression, or upon treatment, in both healthy and diseased cells. A recent study showed that different cell types contribute RNA to the healthy cell-free plasma transcriptome, and that cfRNA allows monitoring of cell-type-specific changes in diseases, such as non-alcoholic fatty liver and Alzheimer’s disease^2^. The potential of cfRNA as prognostic biomarker has also been demonstrated for pre-eclampsia and organ damage in pregnancy^3^. Some studies have also shown potential of using cfRNA biomarkers in certain cancer types^4–6^. Larson et al.^4^ specifically looked at genes that are “dark” (no or few detected transcripts) in plasma from healthy controls but abundant in plasma from lung and/or breast cancer patients and found indications of tissue-of-origin signal. Chen et al.^5^ reported potential of combining human and microbe-derived plasma cfRNA for distinguishing healthy donor and cancer plasma samples (colorectum, stomach, liver, lung, and esophageal cancer) based on profiles of RNAs showing group-level differences. Roskams-Hieter et al.^6^ showed plasma cfRNA classification potential for liver cancer and multiple myeloma patient samples compared to non-cancer controls, with a higher accuracy using learning vector quantization feature selection compared to group-level differential abundance. The authors, however, indicated that liver and bone marrow are major contributors to the plasma transcriptome which may result in more cancer signal.

Here, we further examined the impact of different cancer types on plasma cfRNA profiles in different cohorts (Table 1). We first quantified cell-free mRNAs in different cohorts of blood plasma from cancer patients with locally advanced or metastatic tumors and cancer-free donors to uncover tumor or tissue-of-origin mRNA signatures with diagnostic potential. Results revealed substantial heterogeneity in cell-free mRNA abundance among cohorts, questioning the biomarker potential of cell-free mRNA signatures. However, patient-specific alterations, reflected by the number of cell-free mRNAs with differential abundance in a patient sample compared to a cancer-free control population, enabled to accurately distinguish cancer patients from healthy individuals.

**Table 1:** Cohort overview. Name, number of donors (one sample per donor), biofluid type, library preparation method and application of the cohort in this study. (*) eight control donors overlap with pan-cancer cohort, data from these eight donors was only used for discovery of the tail gene concept, not for replication of pan-cancer analyses (see Methods).

| cohort name | cancer patients<br>(cancer types) | controls | biofluid | library preparation | purpose |
| --- | --- | --- | --- | --- | --- |
| pan-cancer cohort | 200 (25) | 8 | plasma | mRNA capture (Illumina DNA Prep with Enrichment) | discovery |
| three-cancer cohort | 36 (3) | 30* | plasma | mRNA capture (Illumina DNA Prep with Enrichment) | replication pan-cancer analyses & discovery tail gene concept |
| lymphoma cohort | 43 (2) | 22 | plasma | total RNA (SMARTer Stranded Total RNA-Seq Kit v3 - Pico Input Mammalian) | tail gene concept in independent plasma cohort |
| bladder cancer cohort | 12 (1) | 12 | urine | mRNA capture (TruSeq RNA Exome) | tail gene concept in independent urine cohort |

## Results

### A diverse repertoire of cell-free mRNAs in plasma

To characterize the blood plasma cell-free mRNA transcriptome, we collected blood plasma from a pan-cancer discovery cohort consisting of 208 individuals covering 25 different cancer types and aged-matched healthy controls (Fig. 1a, Supplementary Table 1). MRNA capture sequencing of the pan-cancer plasma cohort resulted in 3 to 56M paired-end reads per sample (median 16M) after quality filtering with no significant differences among groups (Kruskal-Wallis test p > 0.05) (Extended Data Fig.1a). On average 84% (95% CI [82.59, 84.66]) of uniquely mapped reads were assigned to protein coding genes and most other reads originated from mitochondrially encoded ribosomal RNAs (Extended Data Fig.1b). Per sample, between 7439 and 13,039 unique mRNAs (median 10,932) were detected with at least 10 counts – we use the term mRNA to refer to all mRNA reads mapping to the same gene (Fig. 1b). All plasma samples had 5263 mRNAs in common and 9908 mRNAs were found in at least 75% of the samples. Of note, we detected more mRNAs in plasma from acute myeloid leukemia (AML) patients compared to plasma from controls and from patients with solid cancers (Kruskal-Wallis p = 6.6E-4, effect size = 0.163, *post hoc* two-sided Wilcoxon rank-sum q < 0.05 for AML vs controls and 16 out of 24 solid cancer types). Although some AML samples were sequenced deeper, sequencing depth alone did not explain the higher number of detected mRNAs (Extended Data Fig.1c). We next determined the mRNA concentration using synthetic RNA spikes that were added to each lysed plasma sample (Fig. 1c). MRNA concentrations ranged from 0.003 to 8.302 ng/ml with an average concentration of 0.486 ng/ml. Despite the higher number of detected mRNAs, the mRNA-concentration in AML plasma was not significantly higher than controls or other cancer types. Moreover, though the largest median mRNA concentration (READ, 0.648 ng/ml) was more than 10 times higher than the smallest median (ANA, 0.063 ng/ml), no significant concentration differences were found among the groups (Kruskal-Wallis test p = 0.0275, effect size = 0.087, with all *post hoc* two-sided Wilcoxon rank-sum q > 0.05). Besides the small sample sizes, this may be related to the considerable cfRNA concentration variability between patients, which is also reported at cfDNA level^7^.

**Fig. 1:**
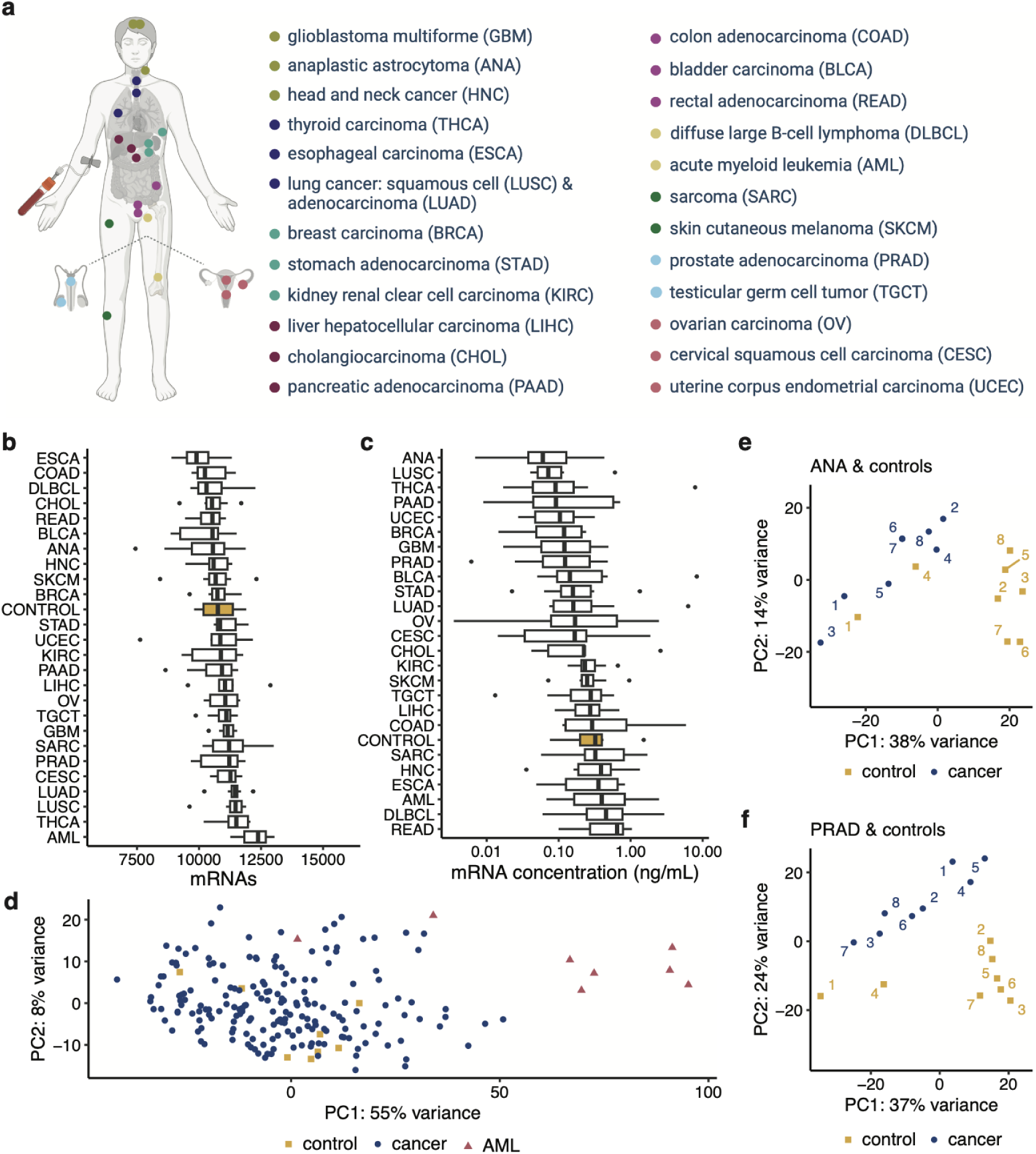
Pan-cancer plasma cohort overview, mRNA content and sample clustering. **a**, Primary sites of the 25 cancer types in the pan-cancer cohort. Primary sites are indicated with colored dots and corresponding cancer types are listed (top to bottom) with the (sub)type abbreviations between brackets. Details can be found in Supplementary Table 1. **b**, Number of plasma mRNAs, detected with at least 10 counts, across cancer types and healthy controls. Groups ordered according to median. **c**, Variability in plasma mRNA concentration across cancer types and healthy controls. Groups ordered according to median. **b** and **c**, Boxplots show lower quartile (Q1), median, and upper quartile (Q3). Whiskers extend from the lower and upper quartile to the smallest and largest value, respectively, within at most 1.5*interquartile range (Q3-Q1) from that quartile. More extreme points are plotted as individual dots. **d**, Principal component analysis using the 500 most variable mRNAs in the pan-cancer cohort (with variance stabilizing transformation DESeq2). Yellow squares: control samples; red triangles: acute myeloid leukemia samples; blue dots: non-leukemia cancer samples. **e** and **f**, Principal component analysis using the 500 most variable mRNAs in controls and anaplastic astrocytoma (ANA) or prostate cancer (PRAD) patients, respectively. Replicate numbers are indicated in the plot. PC: principal component.

We then searched for evidence of tumor-derived cfRNA by investigating known cancer fusion transcripts in individual plasma samples. In every cancer plasma sample, we only considered fusion transcripts that had been documented in tumor tissue of that same cancer type. Such fusion transcripts were only identified in plasma samples of AML patients. The *PML::RARA* fusion transcript^8^ was found in two AML plasma samples with several reads supporting the fusion (7 to 19 spanning pairs and 6 to 9 unique spanning reads). The reciprocal *RARA::PML* fusion was found in one of both AML plasma samples (19 pairs and 17 unique reads). In plasma samples of solid cancer types, we only detected fusion transcripts that had not been reported in the tumor tissue of these cancer types (Supplementary Table 2). Of note, these fusion transcripts had fewer supporting reads (1 to 2 pairs and 2 to 5 unique reads) than the *PML::RARA*/*RARA::PML* fusions in AML and may thus represent false positives. Moreover, other fusions with few supporting reads were also identified in healthy control plasma samples. While the two AML samples with the *PML::RARA* fusion transcripts were sequenced deeper than other samples, the fusions were still detected when downsampling to the median number of reads of the pan-cancer cohort. These analyses confirm the presence of tumor cell-derived cfRNA in plasma samples from AML patients.

### Differential mRNA abundance in cancer patients’ plasma compared to controls

Principal component analysis revealed a clear separation between AML and other plasma samples for six out of eight AML patients, with and without downsampling AML sample reads to the median of the cohort (Fig. 1d & Extended Data Fig.1d). Control samples mostly clustered in between samples of solid tumors. Analysis of individual solid tumor types revealed that most patient samples could be separated from controls in the first two principal components (Fig. 1e and f, Extended Data Fig.2), suggesting that plasma cfRNA profiles of cancer patients differ from that of controls.

Differentially abundant mRNAs were identified in every cancer-control comparison, with numbers varying from 10 to 2583 (median 128) for higher abundant mRNAs and from 89 to 2727 (median 221) for lower abundant mRNAs in cancer compared to control (Fig. 2a). Most differentially abundant mRNAs were found in AML (5310 mRNAs, or 5347 mRNAs when downsampling reads to the median reads in cohort). Uterine cancer (UCEC) and sarcoma (SARC) completed the top 3 with 1154 and 763 mRNAs, respectively. Head and neck (HNC), rectal (READ) and breast cancer (BRCA) had the lowest number of differentially abundant mRNAs (127, 136, and 145 respectively).

**Fig. 2:**
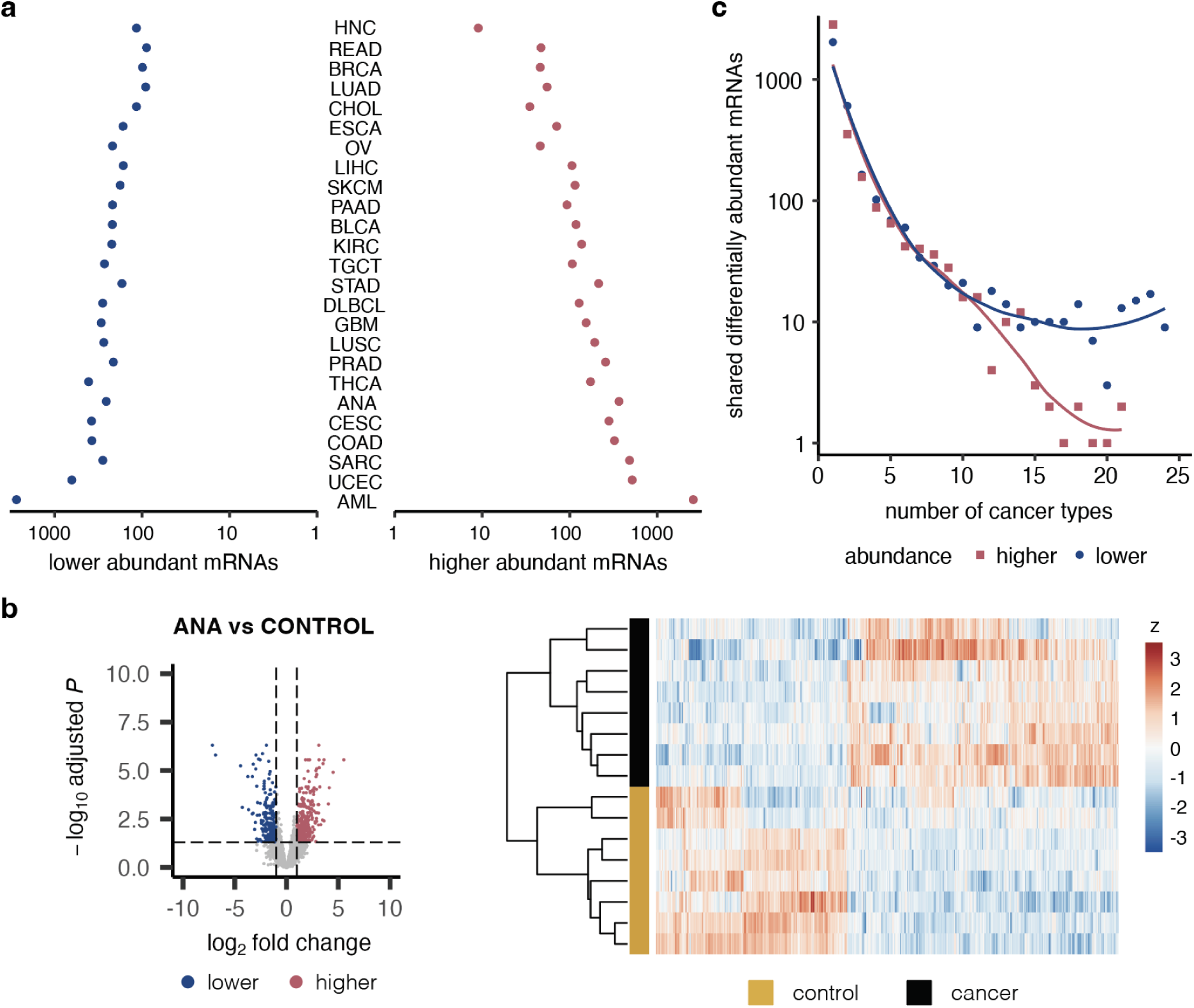
Differentially abundant mRNAs in cancer versus control. **a**, Differentially abundant mRNAs in plasma from cancer patients versus controls. Cancer types ordered based on total number of differentially abundant mRNAs (q < 0.05 and |log_2_ fold change| > 1). **b**, Volcano plot and heatmap of differentially abundant mRNAs in anaplastic astrocytoma vs controls (other cancer-control comparisons in Extended Data Fig.3). Volcano plots have a horizontal line at q = 0.05 and vertical lines at log_2_ fold change of -1 and 1, resp. Higher (red): higher abundant in cancer compared to control samples (q < 0.05 and log_2_ fold change > 1); lower (blue): lower abundant in cancer compared to control samples (q < 0.05 and log_2_ fold change < -1). Heatmaps show relative abundance of differentially abundant mRNAs (columns) in individual plasma samples (rows). Heatmap colors represent log2(normalized counts+1) followed by z-score transformation per gene (z). Clustering of genes and samples based on Pearson correlation. **c**, Recurrence of differentially abundant mRNAs (shown in a) across cancer types.

In total, 6758 unique mRNAs were differentially abundant in one or more cancer-control comparisons (b) and the median absolute fold change was 3.1. One quarter of differentially abundant mRNAs had an absolute fold change of at least 4.6. Hierarchical clustering based on differential mRNA abundance mostly separated cancer and control samples but also revealed variability in gene abundance within groups (Fig. 2b, Extended Data Fig.3).

4442 mRNAs (66%) were differentially abundant in one cancer type only (Fig. 2c), and most (3680 mRNAs) were identified in AML. While many mRNAs were differentially abundant in plasma of only one or a few cancer types, 26 differentially abundant mRNAs were recurrent in 23 out of 25 cancer-control comparisons (Fig.2c). These mRNAs were all lower abundant in cancer compared to controls and showed enrichment for inflammatory and immune response pathways (Supplementary Table 3).

### Enrichment of pathways and cell types in cancer patients’ plasma

To explore the pathways, cell types and tissues underlying the differentially abundant mRNAs in cancer plasma, we first applied gene set enrichment analyses using hallmark^9^, Kyoto Encyclopedia of Genes and Genomes (KEGG)^10^, and custom tissue- and cell type gene sets (Fig.3a, Extended Data Fig.4a & b). We observed a negative enrichment of various immune-related signals (interleukin signaling, interferon gamma and inflammatory response, and neutrophils) across all cancer types except UCEC, with q < 0.05 in at least 21 out of 25 cancer types. We also found a negative enrichment of the KEGG ribosome gene set in all solid cancer types (q < 0.05 in 19 out of 24 types) (Extended Data Fig.4b). These results are in line with a previous study by Chen et al.^5^ on plasma cfRNA profiles of five early-stage solid cancer types. Other negatively enriched gene sets with q < 0.05 in at least half of the cancer types in our cohort were blood related (Fig.3a, Extended Data Fig.4a & b). Conversely, ’extracellular matrix receptor’ and ‘focal adhesion’ gene sets were significantly positively enriched in at least half of the cancer types (Extended Data Fig.4b), confirming results by Chen et al. Other positively enriched gene sets included those related to KRAS signaling, epithelial-mesenchymal transition, myogenesis, adipose tissue, brain function, and hematopoietic stem cells. (Fig.3a, Extended Data Fig.4a).

**Fig. 3:**
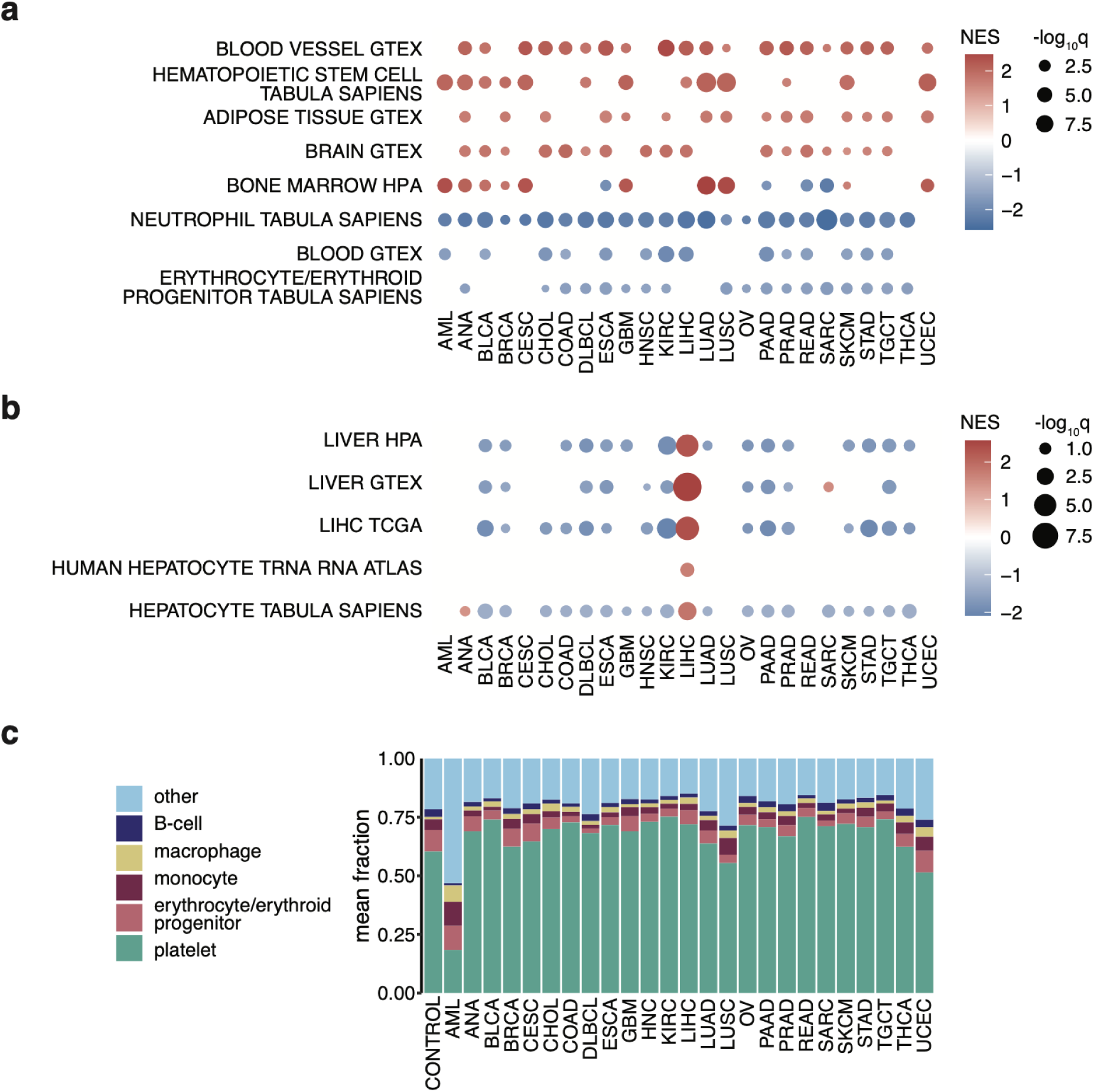
Gene set enrichment and cell type deconvolution reveal pan-cancer and cancer type-specific signals. **a**, Gene set enrichment analysis results for custom tissue and cell type sets. Only sets that are significantly enriched in at least half of the cancer types are shown. Gene sets with q < 0.05 colored according to normalized enrichment scores (NES). **b**, Liver-related gene set enrichment in plasma from liver cancer patients, gene sets with q < 0.25 colored according to NES. Detailed information about gene sets can be found in Methods. **c**, Average fraction of the five most abundant cell types based on deconvolution. Other: cumulative sum of other cell types. The five most abundant cell types were selected based on mean ranking in all groups.

Cancer type-specific enrichment of cfRNA reflecting the primary tumor site was only observed for liver cancer patients. ‘Liver tissue’, ‘liver cancer tissue’, and ‘hepatocyte’ gene sets were all positively enriched in liver cancer patients’ plasma (q < 0.05, Fig.3b) and not in plasma from any other cancer type. A ‘bronchial epithelial cells’ gene set was significantly enriched in lung adenocarcinoma and lung squamous cell carcinoma (LUAD q = 0.027; LUSC q = 0.026), but this enrichment was also found in four other cancer types (ANA, CESC, CHOL, and UCEC).

We then applied computational deconvolution to enumerate the cell type fractions contributing to plasma cfRNA profiles (Supplementary Table 4). Platelets, erythrocytes/erythrocyte progenitors, monocytes, macrophages, and B-cells formed the top-5 of most abundant cell types that contributed to plasma cfRNA of both controls and patients with solid tumors (Fig.3c). Except for macrophages, these cell types were also reported as the most abundant cell types in a previous study on healthy donor plasma^2^. Plasma from AML patients showed a very different composition. Several cell type contributions - including hematopoietic stem cells, myeloid progenitor cells, and monocytes - were increased or even exclusively present in AML plasma samples, in line with the blood cell counts typically observed in these patients^11^. Myeloid progenitor cells, normally not present in peripheral blood, were an abundant contributor in the AML plasma samples (mean 10.7%, or 8.5% when downsampling AML reads to the cohort median), with almost no signal in other plasma samples (mean 0.1%) (Extended Data Fig.4c). Moreover, the platelet fraction in AML plasma was significantly lower compared to other groups (two-sided Wilcoxon rank sum p = 3.8E-5, effect size = 0.289) with on average 18.3% in AML compared to 62.3% in other samples, reflecting the reduced thrombocyte counts commonly found in leukemia patients^11^.

In plasma from patients with solid tumors, the platelet fraction was significantly higher compared to controls (Extended Data Fig.4c, two-sided Wilcoxon rank-sum p = 2.3E-2, effect size = 0.163), which is in line with the prognostic value of platelet counts and the frequent occurrence of thrombocytosis in solid tumor patients^12,13^. For neutrophils, erythrocyte/erythroid progenitor cells, thymocytes, and T-cells, the fraction was significantly lower in cancer patients’ plasma compared to controls (two-sided Wilcoxon rank-sum p = 1.8E-2 (effect size = 0.169), p = 1.4E-2 (effect size=0.176), p = 1.8E-2 (effect size=0.164), p = 4.3E-4 (effect size = 0.252), respectively).

### Independent cohort confirms systemic signal and cfRNA heterogeneity

To verify our findings, we processed an independent (three-cancer) cohort of plasma samples which, after removing samples with low read depth, included samples from 20 cancer-free control donors and 35 cancer patients with one of three different solid cancer types: prostate (PRAD, n = 12), ovarian (OV, n = 11), and uterine (UCEC, n = 12) cancer. 7 to 61M paired-end reads were obtained per sample (median 28M) with no significant difference in sequencing depth among groups (Kruskal-Wallis p = 0.7858) (Extended Data Fig.5a). In line with the pan-cancer cohort, we detected between 8642 and 11,944 mRNAs per sample (Extended Data Fig.5b), with 6194 mRNAs shared across all plasma samples of this three-cancer cohort. MRNA concentrations ranged from 0.007 to 0.414 ng/ml (Extended Data Fig.5c).

Most cancer samples could be separated from controls in the first principal component, especially for prostate and ovarian cancer (Fig.4a). Based on differential abundance analyses between cancer and control samples, we found the highest number of differentially abundant mRNAs in OV (995 higher abundant, 1644 lower abundant), followed by PRAD (848 higher abundant, 1043 lower abundant) (Fig.4b). The lowest number of differentially abundant mRNAs was found for UCEC (243 higher abundant, 349 lower abundant) even though this cancer type had the most differentially abundant mRNAs in the pan-cancer cohort. Hierarchical clustering based on differential mRNA abundance in this three-cancer cohort mostly separated cancer and control samples but again revealed variability in mRNA abundance within groups (Extended Data Fig.5d).

**Fig. 4:**
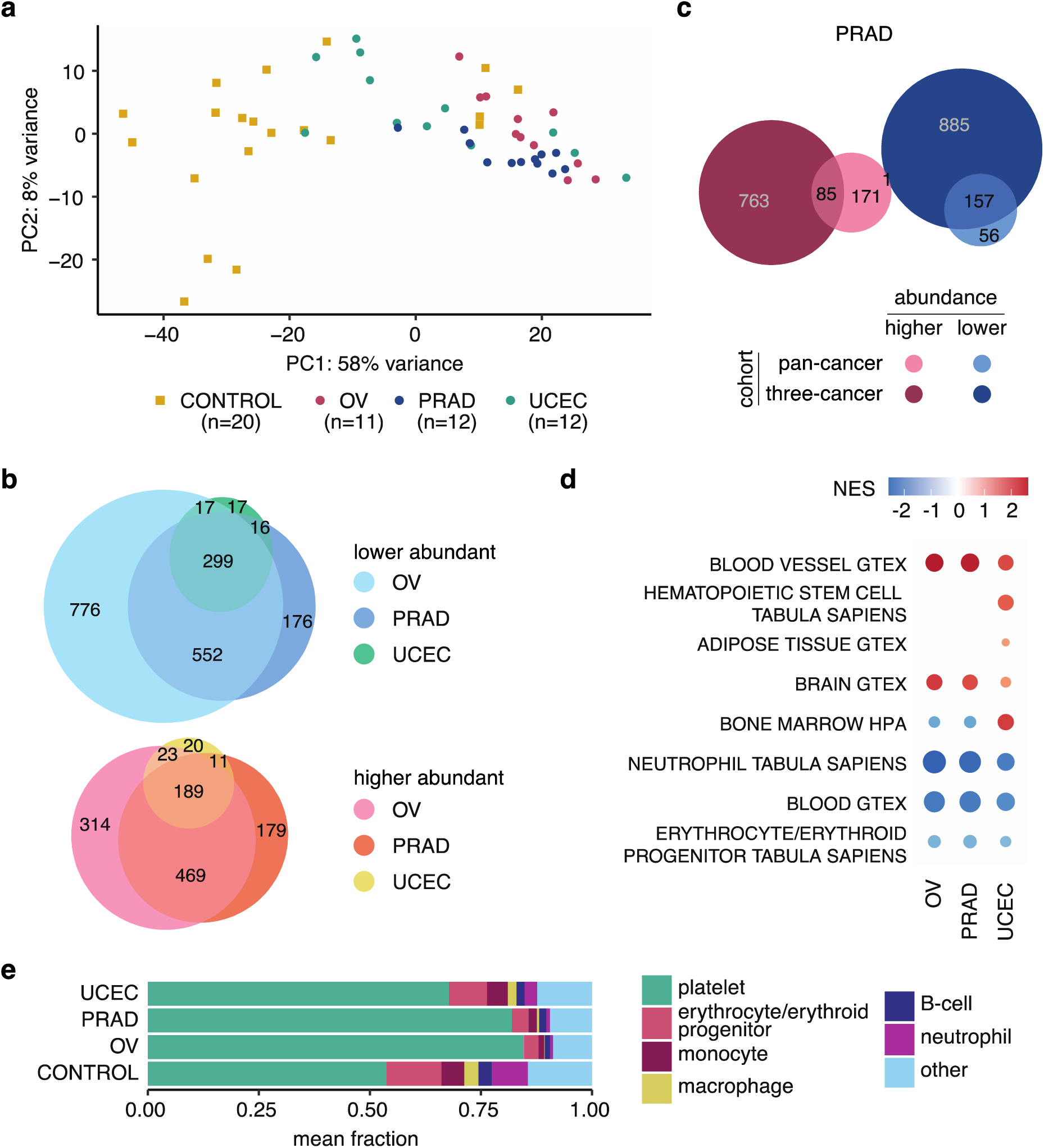
Three-cancer cohort confirms findings from pan-cancer cohort and reveals significant, yet limited, overlap in differentially abundant mRNAs. **a**, Principal component analysis on top 500 most variable mRNAs shows distinct clustering of most cancer and control samples (variance stabilizing transformation DESeq2). **b**, 0verlap between differentially abundant mRNAs detected in individual cancer-control comparisons. Circles are proportional to the number of genes. **c**, Overlap between differentially abundant genes in prostate cancer versus control for the pan-cancer and three-cancer cohort. Fisher’s exact test: p = 2.9E-52, odds ratio = 12.2, Jaccard index = 0.083 for higher abundant mRNAs in cancer versus control (higher); p = 1.1E-155, odds ratio = 59.3, Jaccard index = 0.143 for lower abundant mRNAs (lower). Circles are proportional to the number of genes. **d**, Gene set enrichment analysis results for custom tissue and cell type sets. Only pathways of Fig.3a shown. Gene sets with q < 0.05 colored according to normalized enrichment scores (NES). **e**, Average fraction of the five most abundant cell types and macrophages based on cell type deconvolution. Other: cumulative sum of other cell types. Differential abundance: q < 0.05 & |log_2_ fold change| > 1. OV: ovarian cancer; PRAD: prostate cancer; UCEC: uterine cancer.

189 and 299 mRNAs were significantly higher and lower abundant, respectively, in plasma from all three cancer types compared to controls (Fig.4b). The lower abundant mRNAs included all 26 mRNAs that were identified as lower abundant in most cancers in the pan-cancer cohort. There was a significant, although limited, overlap in differentially abundant mRNAs between the three-cancer and the pan-cancer cohort for each of the three cancer types (Fisher’s exact test on lower abundant mRNAs: OV p = 8.4E-118 (odds ratio = 30.0), PRAD p = 1.1E-155 (odds ratio = 59.3), UCEC p = 7.6E-13 (odds ratio = 4.2); on higher abundant mRNAs: OV p = 4.0E-4 (odds ratio = 4.6), PRAD p = 2.9E-52 (odds ratio = 12.2), UCEC p = 2.2E-5 (odds ratio = 3.3); Jaccard index [0.009-0.143]) (Fig.4c and Extended Data Fig.6a). Despite the uniform collection and processing of samples from patients with the same cancer type, the low Jaccard indices indicate high heterogeneity between the two cohorts, as illustrated with probabilistic modelling for varying cohort sizes (Supplementary Analysis 1). Respectively 38%, 49%, and 95% of the differentially abundant mRNAs in OV, PRAD, and UCEC in the pan-cancer cohort could not be confirmed in the three-cancer cohort. Notably, mRNAs with lower abundance showed a higher overlap between both cohorts than mRNAs with higher abundance.

In line with the pan-cancer cohort, there was no enrichment of cancer type-specific gene sets reflecting tumor tissue of origin in the OV, PRAD and UCEC samples in the three-cancer cohort. However, enriched gene sets shared among at least half of the cancer types in the pan-cancer cohort were confirmed in the three-cancer cohort, except for adipose tissue (Fig.4d, Extended Data Fig.6b and c).

Cell type deconvolution confirmed platelets, erythrocyte/erythroid progenitor cells, monocytes and B-cells among the top-5 cell types contributing cfRNA to plasma (Fig.4e, Supplementary Table 4), with neutrophils being ranked as the fourth most abundant cell type. The higher platelet fraction in cancer patient plasma was even more pronounced in the three-cancer cohort (two-sided Wilcoxon rank-sum cancer versus control p = 5.8E-9, effect size = 0.679), especially for ovarian and prostate cancer (Extended Data Fig.6d). Neutrophils, erythrocyte/erythroid progenitor cells, thymocytes and T-cells again had a lower plasma fraction in cancer patients compared to controls (two-sided Wilcoxon rank-sum p=4.4E-6 (effect size = 0.556), p = 3.2E-7 (effect size = 0.610), p = 6.4E-5 (effect size = 0.419), p = 4.1E-2 (effect size = 0.261), respectively).

Taken together, the limited overlap of differentially abundant mRNAs between the pan-cancer and three-cancer cohort highlights plasma transcriptome variability, yet enrichment and cell type contribution analyses confirm the systemic cfRNA changes in plasma from cancer patients.

### cfRNA tail genes distinguish cancer samples from controls

Apart from liver cancer and AML, our analyses did not reveal evidence for a tumor or tissue of origin signal among mRNAs that were more abundant in cancer patient versus control plasma. Moreover, higher abundant mRNAs were less recurrent across cancer types and showed lower reproducibility between cohorts compared to mRNAs with lower abundance in cancer versus control plasma. Our analyses suggest that the latter represent a systemic immune response rather than a tumor derived signal. Based on these observations, we hypothesized that (tumor derived) cfRNA signals may be heterogeneous among patients, and that classic differential mRNA and pathway abundance analysis between groups may not reveal such non-uniform signals. We therefore decided to investigate cfRNA profiles in individual cancer patients using samples from the three-cancer cohort.

To capture aberrations in the cfRNA profile of an individual cancer patient, we compared the mRNA abundance of a cancer plasma sample to the mRNA abundance distribution in the entire control group, that served as a reference. More specifically, a z-score was calculated for each mRNA based on the mean and standard deviation of the log_2_ normalized count distribution in the control group. Abundant mRNAs with an absolute z-score of at least 3 were defined as “tail genes” for that patient, referring to their position in one of the tails of the control z-score distribution (Fig.5a).

**Fig. 5:**
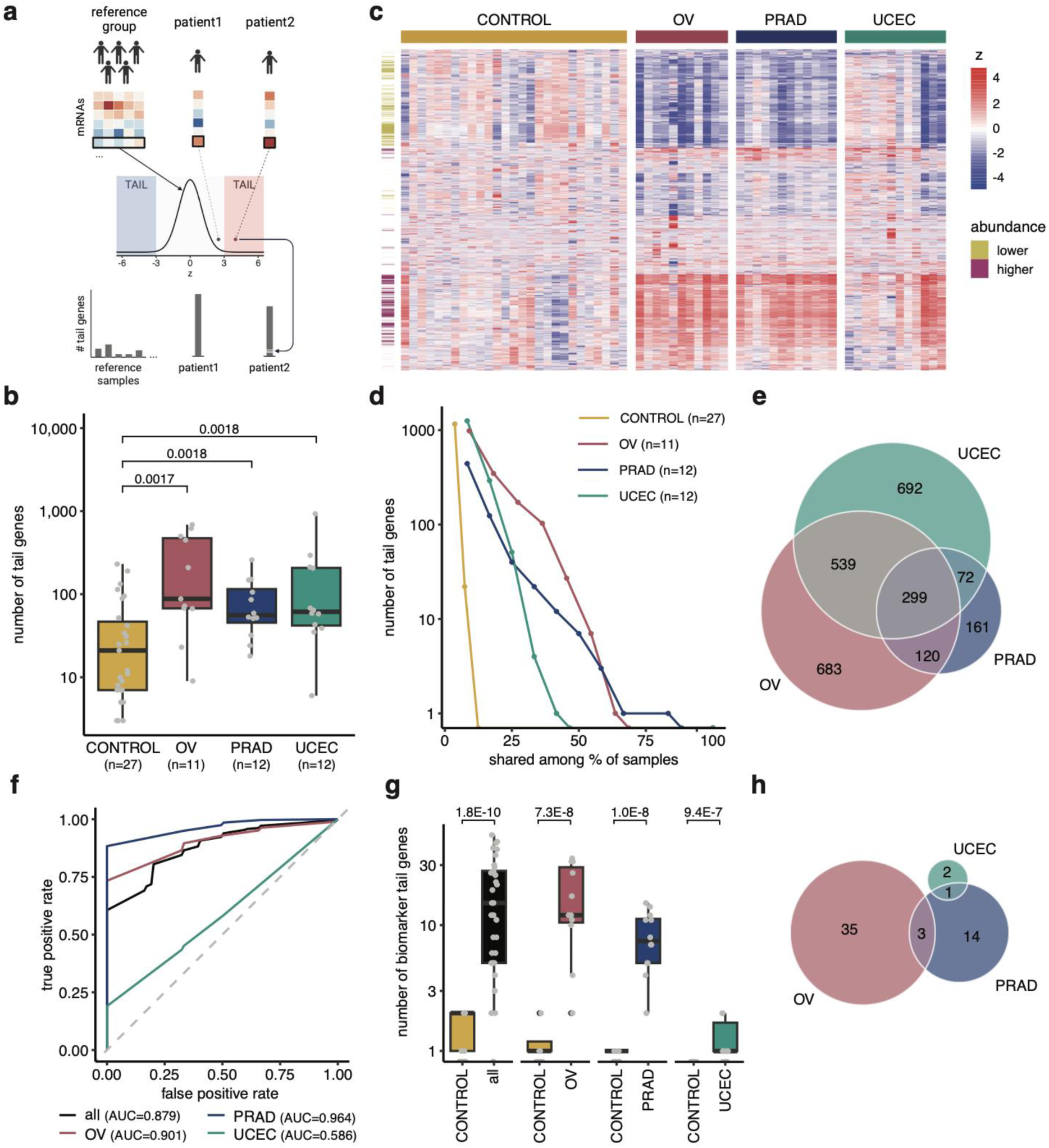
Higher number of (biomarker) tail genes in cancer patient plasma samples compared to controls. **a**, Tail gene identification workflow. A gene in a particular sample for which the abundance deviates more than 3 standard deviations from the reference group abundance distribution is a tail gene for that sample. The number of tail genes is used as a metric to determine cancer/control status. **b**, Boxplot of the number of tail genes per group, with grey dots representing individual sample counts. Kruskal-Wallis p = 0.001, effect size = 0.222. Two-sided Wilcoxon rank-sum q-values for specific cancer versus control comparisons indicated in the plot. **c**, Z-score distribution of tail genes in the different samples, all 3312 genes that fulfilled tail gene criteria in at least one of the samples are shown. For visualization purposes, z-scores above 5 or below -5 were reduced to 5 and -5, respectively. Genes (rows) clustered based on Euclidean distance. **d**, Number of tail genes shared by a certain fraction of samples of the same type. Number of samples indicated between brackets. **e**, Overlap between tail genes detected in the different cancer types. **f**, mean ROC curves of 10x 5-fold cross-validation for binary cancer/control classification in test sets based on the number of biomarker tail genes for control and ovarian cancer samples (OV), prostate cancer samples (PRAD), uterine cancer samples (UCEC), or all cancer samples (all) using the respective biomarker tail gene sets in the training sets. **g**, Boxplot of number of 50% consensus biomarker tail genes per subset in control (CONTROL) versus the respective cancer samples. Total number of biomarker tail genes considered for each comparison: 63 for all cancer (n = 35) versus control, 38 for OV versus control, 18 for PRAD versus control, 3 for UCEC versus control. Grey dots represent individual sample counts. Two-sided Wilcoxon rank-sum p-values indicated in plot. **h**, Overlap of cancer type-specific biomarker tail genes. Boxplots show lower quartile (Q1), median, and upper quartile (Q3). Whiskers extend from the lower and upper quartile to the smallest and largest value, respectively, within at most 1.5*interquartile range (Q3-Q1) from that quartile. More extreme points are plotted as individual black dots. 62 plasma samples considered in total from 27 cancer-free controls (CONTROL), 11 ovarian cancer (OV), 12 prostate cancer (PRAD), and 12 uterine cancer patients (UCEC)).

We searched for tail genes in each cancer patient from the three-cancer cohort (where we could take advantage of a large control group) and in each control sample, always using all other control samples as reference (Extended Data Fig.7a & b). We identified a total of 3311 unique tail genes in this cohort and observed a significantly higher number of tail genes in plasma samples from cancer patients compared to controls (Kruskal-Wallis p = 0.0012, effect size = 0.222, Fig.5b). 74% of the tail genes were not identified as differentially abundant in cancer samples versus controls (Fig.5c). Moreover, for 40.0% of cancer sample tail genes that were differentially abundant between cancer and controls, the direction (i.e. positive or negative z-score) in at least one cancer sample was opposite to the direction reported by the differential abundance analysis, further highlighting the heterogeneity among patients. Notably, the recurrence of tail genes was much higher in cancer samples than controls; 98% of the control tail genes were identified in only one control sample while 47% of cancer tail genes were identified in more than one cancer sample (Fig.5d).

To determine the significance of the observed results, we randomly swapped sample labels prior to tail gene identification and repeated this procedure 20 times. Doing so, we did not observe a significant difference in number of tail genes between the newly labeled cancer samples and controls in any of the repeats (all Kruskal-Wallis p≥0.05), confirming that an increased number of tail genes is specifically associated with cancer samples.

Tail genes detected in one cancer type showed significant overlap with tail genes detected in another cancer type (Fisher’s exact tests p≤5.4E-168, odds ratio: [11.1-15.2]), yet limited similarity (Jaccard index: [0.197-0.348]) (Fig.5e). The 1182 unique control tail genes also showed significant overlap with any of the cancer tail gene subsets (pairwise Fisher exact tests p: [5.9E-15 - 8.5E-32], odds ratio: [2.1-2.5]) with less similarity (Jaccard index: [0.074-0.119]). 63% of control tail genes were not identified as tail gene in any of the cancer samples.

Seven Cancer Gene Census oncogenes were identified as tail genes (z > 3) in at least one sample, which is significantly more than expected by chance (Fisher’s exact p = 0.019, odds ratio = 3.13). While most oncogenes were only identified as tail gene in a single sample, *KAT6A* was found in seven cancer samples (4 OV, 2 PRAD, and 1 UCEC) but also in one control sample. Overrepresentation analyses (Extended Data Fig.8, Supplementary Table 5) demonstrated that tail genes that are higher abundant in at least one cancer sample (z > 3) were involved in different processes including platelet activation and signaling, actin binding and dynamics, and extracellular matrix. Tail genes that are lower abundant in at least one cancer sample (z < -3) were mainly related to mRNA splicing and translation. Significantly enriched processes in the cancer tail gene sets were not enriched in the control tail gene set, and processes enriched in the lower abundant (cancer) tail genes showed higher recurrence across the three cancer types compared to those enriched in the higher abundant tail genes.

Given the higher number of tail genes in cancer versus controls, we further assessed the binary (cancer/control) classification potential of the number of tail genes per sample. We used a 10x 5-fold cross-validation in which we first selected a subset of tail genes, further coined biomarker tail genes (BTG), that were significantly associated to either the cancer or control state using Fisher’s exact tests on the training set (see Methods). This resulted in sets of 43 to 549 BTG. None of these BTG were recurrent tail genes in the control samples, and for BTG identified in at least half of the cross-validation folds, 43% were not identified as differentially abundant in any cancer type compared to controls (Extended Data Fig.9). Applying the same biomarker tail gene selection strategy to 10 iterations of randomly swapping sample labels resulted in considerably fewer or no biomarker tail genes at all: between 0 and 6 BTG (median: 0) instead of 43 to 549.

When selecting biomarker tail genes in a specific cancer group versus controls, we obtained 17 to 271 biomarker tail genes for OV, 6 to 82 for PRAD, and 2 to 16 for UCEC. We then built a classifier for discriminating cancer and control samples based on the number of tail genes belonging to the BTG set in a particular sample. Using 10x 5-fold cross-validation, we obtained a high binary classification for ovarian or prostate cancer samples and controls (mean AUC = 0.901 and 0.964, resp.), yet a lower classification performance for uterine cancer samples and controls (mean AUC = 0.586) (Fig.5f). Classification performance was still high when combining cancer samples for BTG identification (mean AUC = 0.879), with a better performance for male donors compared to female donors (Supplementary Table 6).

The consensus set of BTG present in at least half of the 10x 5-fold cross-validations, consisted of 38 BTG for OV, 18 for PRAD, 3 for UCEC, and 63 for all three cancer types combined. OV and UCEC consensus BTG sets do not overlap. The overlap for OV and PRAD as well as for PRAD and UCEC is significant but the similarity is limited (Fisher’s exact p ≤ 1.6E-2; odds ratio: 18.5 and 94.7; Jaccard index: 0.06 and 0.05) (Fig.5h).

The difference in number of consensus biomarker tail genes between cancer and control samples per biomarker tail gene subset was more pronounced than based on all tail genes (two-sided Wilcoxon rank-sum: for OV p = 7.3E-8 and effect size = 0.876; for PRAD p = 1.0E-8 and effect size = 0.920; for UCEC p = 9.4E-5 and effect size = 0.629; for all cancers combined p = 1.8E-10 and effect size = 0.810) (Fig.5b and g).

Together, these analyses suggest that individual cancer samples can be distinguished from controls based on the number of biomarker tail genes in their plasma cfRNA profiles.

To independently assess the classification potential of the number of biomarker tail genes, we generated additional plasma cfRNA profiles (lymphoma cohort), including plasma samples from 22 cancer-free control donors and 43 lymphoma patients, more specifically 30 diffuse large B-cell lymphoma (DLBCL) and 13 primary mediastinal large B-cell lymphoma (PMBCL) patients. We identified 5374 unique tail genes, again with more tail genes in cancer samples compared to controls (Kruskal-Wallis p = 2.9E-5, effect size = 0.305, Extended Data Fig.10a). All control tail genes were identified in only one control sample while 45.9% of DLBCL and 44.2% of PMBCL tail genes were identified in more than one DLBCL and PMBCL sample, respectively (Extended Data Fig.10b). When swapping sample labels (20 repeats) prior to tail gene identification, no significant differences in the number of tail genes between cancer and control samples were observed in any of the repeats (Kruskal-Wallis p≥0.05).

Among the tail genes, 64 to 326 lymphoma BTG, 21 to 220 DLBCL BTG, and 87 to 677 PMBCL BTG were identified using Fisher’s exact tests in 10x 5-fold cross-validation. The binary classifiers (cancer/control) based on biomarker tail genes in the training sets resulted in a mean AUC of 0.765 for DLBCL, 0.951 for PMBCL, and 0.866 for all lymphoma in the test sets (Fig.6a). Again, we observed a better performance for samples from male compared to female donors (Supplementary Table 6). When looking at biomarker tail genes that were identified in at least half of the iterations, we found 60 consensus BTG for DLBCL, 371 for PMBCL, and 151 for lymphoma. The number of tail genes belonging to these sets was again significantly higher in cancer compared to control samples (Fig.6b). 59% of these consensus BTG were not differentially abundant in any lymphoma subtype versus controls. Notably, early-stage lymphoma patients also displayed an increased number of biomarker tail genes (Fig.6c) and the number of detected BTG per sample was not significantly linked to cancer staging (Jonckheere-Terpstra p = 0.9374 for DLBCL and p = 0.8949 for PMBCL).

**Fig. 6:**
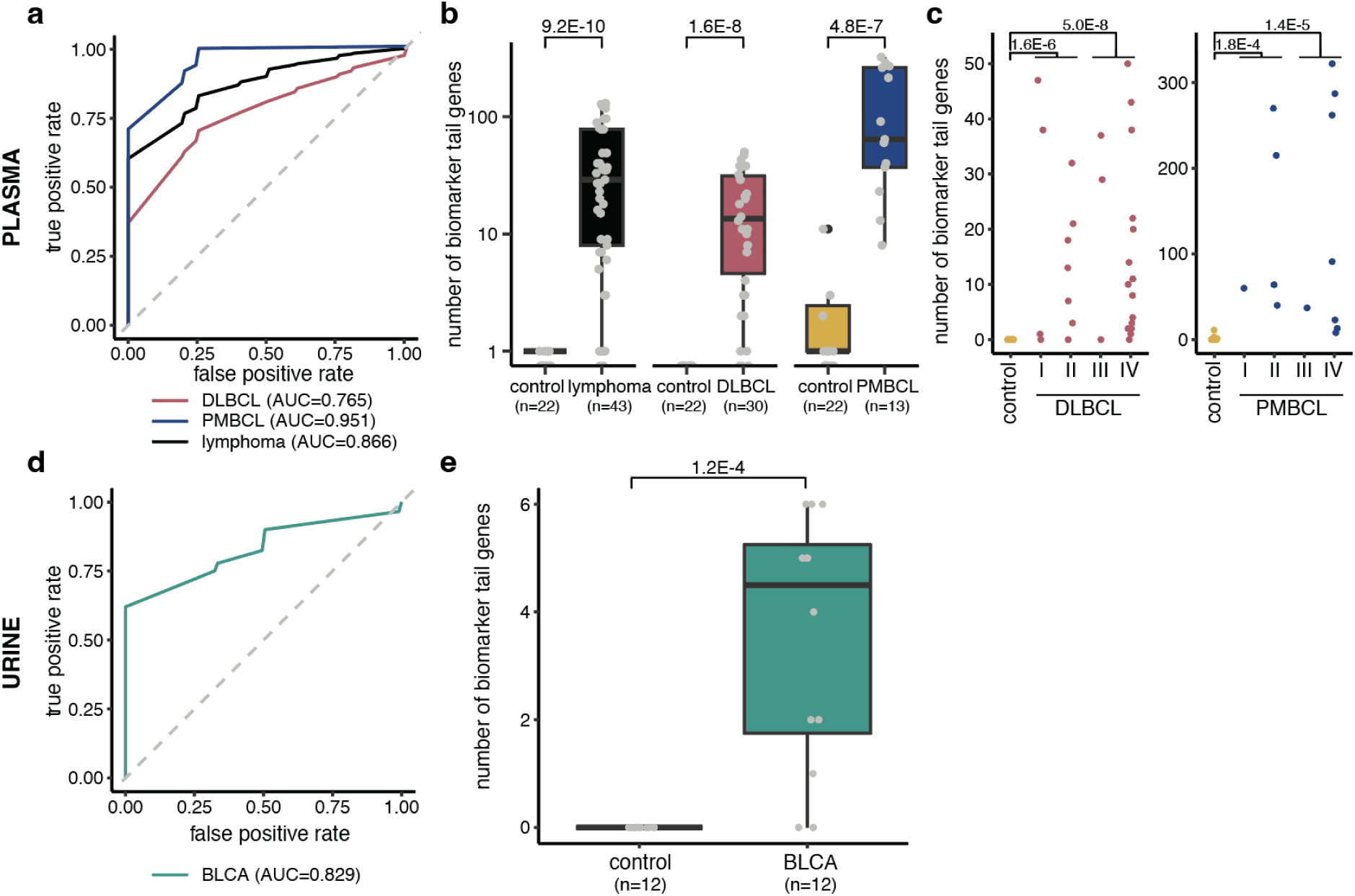
Number of biomarker tail genes is significantly higher in cancer versus control samples in an independent blood plasma and urine cohort. **a-c**, lymphoma cohort: plasma samples from 22 control, 30 diffuse large B-cell lymphoma (DLBCL), and 13 primary mediastinal large B-cell lymphoma (PMBCL) patients. **d-e**, bladder cancer cohort: urine samples from 12 control (CONTROL) and 12 bladder cancer (BLCA) patients. **a**, mean ROC curve of 10x 5-fold cross-validation for binary cancer/control classification based on the number of biomarker tail genes for control and DLBCL, or control and PMBCL using the respective biomarker tail gene sets. **b**, boxplot of number of 50% consensus biomarker tail genes in control versus the respective cancer samples. Total number of biomarker tail genes considered for each comparison: 60 for DLBCL versus control, 371 for PMBCL versus control, 151 for both lymphoma types versus control. Grey dots represent individual sample counts. **c**, number of biomarker tail genes of panel b visualized per cancer stage**d**, mean ROC curve of 10x 5-fold cross-validation for binary cancer/control classification based on the number of biomarker tail genes. **e**, boxplot of number of 50% consensus biomarker tail genes in control versus cancer samples. 6 biomarker tail genes considered in total. Wilcoxon rank-sum p-values indicated in the plots. Boxplots: lower quartile, median, upper quartile, whiskers of 1.5*interquartile range; more extreme points are indicated by black dots.

To test if our cancer-control classification strategy based on biomarker tail genes is also applicable to other biofluids, we applied it to a cohort of urine cfRNA profiles (bladder cancer cohort)^14^. This cohort included urine samples of 12 bladder cancer patients (BLCA) and 12 cancer-free control donors. We identified 1152 tail genes in total and again only observed recurrence in cancer samples (Extended Data Fig.10d). 10x 5-fold cross-validation yielded 2 to 93 biomarker tail genes and the binary classifiers based on these biomarker tail gene sets resulted in a mean AUC of 0.829 in the test sets (Fig.6d). 6 biomarker tail genes were present in at least half of the iterations and the number of tail genes belonging to this 50% consensus set was significantly higher in bladder cancer versus control samples (Fig.6e). In line with the other datasets, sample label swapping prior to tail gene identification did not result in significant differences in the number of tail genes between the newly labeled cancer samples and controls.

Taken together, our results confirm that the number of biomarker tail genes in blood plasma cfRNA enables accurate classification of cancer patients from controls and suggest that the concept may also be applicable to cfRNA profiles from other biofluids.

## Discussion

In this study, we explored cancer-induced changes in blood plasma cfRNA profiles and investigated if these changes could be exploited for classification purposes. Results from our pan-cancer discovery cohort revealed that AML was the only cancer type for which there is direct evidence for tumor-derived cfRNA in plasma in the form of *PML::RARA* fusion transcripts. Also, deconvolution of AML plasma cfRNA revealed a high fraction of myeloid progenitor cells, providing additional – albeit indirect – evidence for tumor-derived cfRNA. For solid tumor types, we identified differentially abundant mRNAs in plasma for every cancer versus control comparison, but a cancer type-specific signal was only observed in plasma from liver cancer patients through the enrichment of liver-specific gene sets. Liver is a major contributor to the plasma transcriptome^4^ and its high vascularization and blood flow may facilitate mRNA release in blood upon liver cancer progression. Increased cfDNA contributions from the liver have also been observed in liver cancer patients^15^. Note that, despite the enrichment of liver-related mRNAs, the total cfRNA concentration was not increased in plasma from liver cancer patients, indicating that this signal was still strongly diluted.

Considering the organ contribution and dilution argument, other types of liquid biopsies may be better suited to monitor tumor-derived cfRNA for other cancer types. For instance, we could not detect enrichment of prostate tissue-related nor prostate cancer-specific transcripts in blood plasma from prostate cancer patients, in contrast to earlier findings in seminal plasma samples^16^.

We did observe several general trends in patients’ blood plasma transcriptomes in the pan-cancer cohort, which were confirmed in an independent cohort. The lower abundance of immune-related mRNAs may be linked to the suppressed immune system status in cancer patients. It was recently shown that local tumors can have a systemic impact on the immune system, and that different tumor sites of origin resulted in distinct relative abundances and activity profiles of immune cell types^17^. This may be linked to the observed variations in immune-related mRNA abundance, both between and within cancer types. The (immune) cell-related mRNA contributions agreed with reported cell abundance trends found in blood from several cancer patients^18^, with the exception of neutrophil-related mRNAs that are lower abundant in plasma. However, neutrophils can have a dual role in cancer^19^ and reduced neutrophil cell-free mRNA abundance may result from neutrophils specifically recruited to tumor sites. Finally, the negative enrichment of blood/erythrocyte gene sets may be linked to anemia, a common condition in cancer patients. Among the processes that we found positively enriched in cancer patient plasma, some are linked to hallmarks for cancer progression and metastasis, including epithelial-to-mesenchymal transition and increased blood vessel signal that stem from leaky vasculature and angiogenesis^20^.

A limitation of the pan-cancer cohort is that control samples were not always perfectly age and sex-matched. The anaplastic astrocytoma, cervical cancer, and testicular germ cell tumor patients were younger as these types predominantly appear at younger age. A dedicated cohort with matched control samples may be better for revealing cancer-related changes in these cancer types. Similarly, while male and female donors were balanced in the control group, this was not always the case for some cancer types due to sex-specificity (e.g. prostate, testicular, breast, cervical, and ovarian cancer) or sample availability. Nevertheless, at most 4 out of 8 donors were not sex-matched in cancer vs control group comparisons in the pan-cancer cohort, and our main conclusions were not impacted when only using controls of the same sex in the three-cancer cohort. Another limitation is the relatively small sample size per cancer type, which might contribute to false positive results or insufficient power to detect true differences.

The variable mRNA abundance in plasma, combined with the limited overlap of differentially abundant mRNAs across cohorts of the same cancer type prompted an alternative approach that can effectively deal with patient heterogeneity. The tail gene concept satisfies this requirement by identifying mRNAs whose abundance in individual samples significantly deviates from the abundance distribution in a reference group. While the dark channel biomarker gene concept^4^ is in part based on a more patient-centric approach, it also requires differential abundance at group level to be considered as potential biomarkers. Moreover, the dark channel concept looks explicitly at genes that are absent (dark) in control samples and therefore cannot capture changes in transcript abundance of genes that are also present in healthy plasma transcriptomes. Of note, 19 dark channel biomarker genes were identified for lung and breast cancer by Larson et al.^4^ but five of those genes could be readily detected in control samples from our cohorts, indicating that the “darkness” (i.e. absence of gene transcripts in control samples) should be re-evaluated in different cohorts.

Tail gene counts are a simple metric for classifying a biofluid sample as originating from a cancer patient or control. Considering that it is not based on gene signatures with weighted contributions of individual features, we propose it is more robust and compatible with observed patient heterogeneity. Nevertheless, it requires a large and representative group of cancer-free control samples. A control group only needs to be established once, if the entire workflow – from sample collection to RNA-sequencing – is standardized across cancer and control samples. This approach should minimize the introduction of batch effects that may skew cfRNA distributions between a pre-established cohort of cancer-free controls and newly profiled cancer or control samples. In this regard, our lab has demonstrated that preanalytical variables (blood collection tube, time between blood collection and plasma preparation, RNA extraction kit) can indeed dramatically impact cfRNA profiles^21^.

Deviations in the number of tail genes are not necessarily restricted to cancer patients, nor to blood plasma. Indeed, cfRNA from urine provides a good source of biomarker tail genes to identify bladder cancer patients. Some of the identified biomarker tail genes have previously been linked to cancer, with examples provided in Supplementary Analysis 3. Notably, other diseases may also be characterized by an increase in tail genes, though the specific (biomarker) tail genes may differ.

Given the limited sample sizes, we likely did not capture all variability in the current tail gene sets, nor did we identify all possible tail genes. Follow-up studies with larger cohorts are recommended to establish robust gene sets. Also, the applicability and overlap of tail genes in non-malignant disease cohorts needs to be examined further. Similarly, while we have preliminary evidence that biomarker tail genes are also significantly more numerous in low stage cancer patients, additional follow-up studies comprising different stages across various cancer types could reveal whether the concept is suited for early-stage cancer screening or monitoring.

## Methods

### Sample cohorts

The study was approved by the ethics committee of Ghent University Hospital, Belgium (no. B670201734362) and performed in accordance with the Declaration of Helsinki.

### Pan-cancer plasma and three-cancer cohort

Blood plasma samples of 266 human donors were obtained from Proteogenex (CA, USA) with informed consent from all donors. The pan-cancer discovery cohort covers 25 different cancer types and a cancer-free control group. For each type, plasma was collected from 8 different donors. The three-cancer cohort consisted of prostate cancer (PRAD), ovarian cancer (OV), and endometrial cancer (UCEC) plasma samples, with 12 donors per type. This cohort also included plasma samples from the 8 control donors of the pan-cancer cohort as well as from 22 additional control donors. The control donors in both cohorts where volunteers without (diagnosed) malignant disease. For independently checking reproducibility of pan-cancer results, only the control samples from the additional control donors were used. For tail gene discovery, all control samples in the three-cancer cohort were used.

All cancer patients in both cohorts had locally advanced (stage 3) to metastatic cancer (stage 4), except for 6 seminoma patients (stage 1). Blood was drawn before treatment with 6 exceptions: 4 glioblastoma patients (grade 4-5) already received chemo and radiation therapy, one prostate cancer patient with metastasis received hormone therapy, and one melanoma patient with metastasis received immunotherapy. Demographic and clinical information can be found in Supplementary Table 1.

### Lymphoma cohort

The lymphoma cohort consisted of plasma samples from 65 human donors: 30 DLBCL patients, 13 PMBCL patients, and 22 control donors. The control donors were volunteers without (diagnosed) malignant disease and without indications of abnormalities in routine blood analyses in the clinical lab. Plasma samples were collected following approval by the ethics committee of Ghent University Hospital, Belgium (B670201733701), and written, informed consent was obtained from all patients.

### Bladder cancer cohort

The bladder cancer cohort is described in more detail elsewhere^14^. This cohort consists of urine samples from 12 muscle-invasive bladder cancer patients and 12 control donors.

### Blood collection and plasma preparation

Blood from the pan-cancer and three-cancer cohort donors was collected in EDTA vacutainer tubes (Becton Dickinson, 367525). Blood was stored at 4 °C until plasma preparation which started by gently inverting the EDTA tube 10 times and centrifugating for 10 min at 1500 g (without brake at 4 °C). Supernatans was then transferred to 15 mL centrifuge tubes and centrifuged again for 10 min at 1500 g (without brake at 4 °C). The resulting platelet depleted plasma was transferred to 2 mL cryovials, frozen and stored at -80 °C within 4 hours of blood collection. All plasma preparations were done in the same lab. Plasma samples were then shipped on dry ice and only thawed on ice immediately before RNA isolation.

For the lymphoma cohort, 2.5 mL of blood was collected from each donor in PAXgene Blood DNA Tubes (BD Biosciences, 761165). Plasma was prepared in one lab by a one-step 15 min centrifugation at 1900 g without brake (room temperature) and stored at -80 °C within 4 hours of blood collection.

### RNA isolation and gDNA removal

RNA isolation and subsequent library preparation was done by one lab technician per cohort. RNA was isolated from 200 µL of plasma with the miRNeasy Serum/Plasma kit (Qiagen, 217184), according to the manufacturer’s instructions. For the pan-cancer and three-cancer cohort, 2 µL Sequin spike-in controls (1/1,300,000 of stock solutions mix A, Garvan Institute of Medical Research, Australia) was added to the lysate during RNA isolation as previously described^22^. For the lymphoma cohort, 2 µL of Sequin spike-in controls were added to the sample lysates (1/5000 of stock solution mix A (Garvan Institute of Medical Research) and 2 µL of External RNA Control Consortium (ERCC) spike-in controls (ThermoFisher Scientific, 4456740) were added to 12 µl RNA eluate.

Genomic DNA was removed by adding 1 μL HL-dsDNase (ArcticZymes, 70800-202) and 1.6 µL reaction buffer (ArcticZymes, 66001) to 12 µL RNA eluate, 10 min incubation at 37 °C, followed by 5 min incubation at 55 °C. RNA was stored at -80 °C and thawed on ice immediately before library preparation.

### Messenger RNA capture library preparation, sequencing, and quantification

MRNA capture library preparation in pan-cancer and three-cancer cohorts started from 8.5 µL DNase treated RNA eluate. cDNA synthesis was performed using TruSeq RNA Library Prep for Enrichment (Illumina, 20020189) as previously described^22^. Briefly, RNA was fragmented, and first strand cDNA was generated using random priming. RNA templates were subsequently removed and replaced by a newly synthesized second strand of cDNA. AMPure XP beads (Beckman Coulter Life Sciences, A63881) were used for purifying the blunt-ended double stranded cDNA. 30 μL cDNA of each sample was then used as input for Illumina DNA Prep with Enrichment (previously Nextera Flex for Enrichment; Illumina, 20025524). Tagmentation and amplification steps were done according to manufacturer’s instructions. Quality of resulting pre-enriched libraries was assessed using a high sensitivity Small DNA Fragment Analysis Kit (Agilent Technologies, DNF-477-0500). The libraries were randomly pooled per 8 or 6 samples, for the pan-cancer and three-cancer cohort, respectively, based on their relative concentrations determined using qPCR for reference genes YWHAZ, ACTB, B2M, and UBC. Each multiplex pool was concentrated to a volume of 15 µL with AMPure XP beads (Beckman Coulter Life Sciences, A63881). Finally, enrichment was performed using probes from the Illumina Exome Panel (Illumina, 20020183), probes complementary to the spike-in controls, and blocking probes against globin (anti-CEX) as previously described^22^. At the end of the enrichment workflow, equimolar library pools were prepared based on qPCR quantification with KAPA Library Quantification Kit (Roche Diagnostics, KK4854).

Paired-end sequencing of both cohorts was performed (2x100 nucleotides) on a NovaSeq 6000. For the pan-cancer cohort, a NovaSeq S2 kit (Illumina, 20028315) was used with standard workflow loading of 1.55 nM (1% PhiX). For the three-cancer cohort, a NovaSeq S1 kit (Illumina, 20028318) was used with Xp workflow loading of 1.25 nM (2% PhiX).

Adapter trimming and removal of reads shorter than 20 nucleotides was done with cutadapt (v1.18). Read quality was assessed with FastQC (v0.11.9) and low-quality reads were filtered: only reads where at least 80% of bases in both mates have a quality score ≥20 (99% accuracy) were kept (between 87 and 99% of reads/sample). Samples with very few reads (< 2M) were removed from all analyses. More specifically, 4 samples were removed in the pan-cancer plasma cohort (1 lymphoma, 1 bladder, 1 head and neck, and 1 lung cancer sample) and 4 samples in the three-cancer cohort (3 control samples, of which 1 repeated from first cohort, and 1 ovarian cancer) as indicated in Supplementary Table 1. Quality filtered reads were then mapped with STAR^23^ (v2.6.0) using the default parameters (except for --twopassMode Basic, --outFilterMatchNmin 20 and --outSAMprimaryFlag AllBestScore). The reference files for all analyses were based on genome build GRCh38 and transcriptome build Ensembl v91, complemented with spike annotations. Finally, gene counts were determined with HTSeq^24^ (v0.11.0) in non-stranded mode, only considering uniquely mapping reads.

### Total RNA library preparation, sequencing, and quantification

Total RNA sequencing libraries of the lymphoma cohort were prepared starting from 8 µL of RNA eluate using the SMARTer Stranded Total RNA-Seq Kit v3 – Pico Input Mammalian (Takara, 634487) according to the manufacturer’s protocol. Equimolar library pools were prepared based on qPCR quantification with KAPA Library Quantification Kit (Roche Diagnostics, KK4854). The libraries were paired-end sequenced (2x100 nucleotides) on a NovaSeq 6000 instrument using a NovaSeq S2 kit (Illumina, 20028315) with standard workflow loading of 0.65 nM (2% PhiX).

After adapter trimming with cutadapt (v1.18) and quality control with FASTQC (v0.11.9), reads were mapped with STAR^23^ (v2.7.3) using default options and the GRCh38 reference described above. Resulting BAM files were deduplicated using UMI-tools (v1.0.0) based on the unique molecular identifier (UMI) sequences in the Pico v3 SMART UMI adapters. Gene counts were determined with HTSeq^24^ (v0.11.0) in reverse stranded mode, only considering uniquely mapping reads.

### DNA contamination check

To assess possible DNA contamination, we looked at splicing and exonic coverage. For each cohort, the number of reads going to spliced junctions was determined using samtools (v1.16.1) and regional coverage was determined using a combination of samtools (v1.16.1) and bedtools (v2.30.0) with BED files containing intronic, intergenic and exonic regions. The summary of splicing and regional coverage results can be found in Supplementary Table 8.

### Data analyses

Most data processing and visualization was done in R (v4.2.1) using tidyverse (v1.3.2) and pheatmap (v1.0.12). Other tools and R packages used for specific analyses are mentioned below. Color schemes are based on Paul Tol’s technical note^25^.

### Detection threshold

To remove noisy datapoints, we applied a threshold of at least 10 counts. This threshold was based on the median threshold that removes at least 95% of single positive genes^26^ - genes that have counts in one replicate but not in the other - between control samples.

### Spike counts and mRNA concentration

Spike counts and Spearman correlations are shown in Supplementary Analysis 3. MRNA concentration was determined based on spikes as previously described^22^. Briefly, the mass of a specific Sequin spike-in control was calculated based on the input concentration and volume of spike-in mix added to the sample. The corresponding mRNA concentration was then estimated by multiplying the spike mass by the ratio of reads mapped to the human genome versus the number of reads mapped to the specific spike, and finally dividing the obtained mass by the plasma volume of the sample. In this study, all Sequin spikes above detection threshold were considered and the geometric mean per sample was taken as the sample’s mRNA concentration.

### Fusion gene detection

FusionCatcher^27^ (v1.30) was used with default settings for fusion transcript identification. Stringent filtering was applied to exclude potential false positives: transcripts with a FusionCatcher label indicative for false positives, transcripts with reads simultaneously mapping to both fusion partners, and transcripts with fusion partners less than 100 kbp apart were filtered out. Only exon-exon fusions were included and detected fusions in plasma were compared to fusions in the Fusion Gene annotation DataBase (FusionGDB)^28^, focusing on fusion genes identified in tissue samples of The Cancer Genome Atlas^29^.

### Differential abundance and principal component analysis

DESeq2 (v1.36.0) was used for normalization and differential abundance analysis. MRNAs were pre-filtered by requiring gene counts equal to or greater than the detection threshold in at least half of the tumor or half of the control samples. In the DESeq2 result table, genes with a Benjamini-Hochberg corrected p-value (q) below 0.05 and an absolute log_2_ fold change above 1 were considered differentially abundant in cancer versus control (higher abundant: q < 0.05 and log2 fold change > 1; lower abundant: q < 0.05 and log2 fold change < -1). Principal component analysis was performed with DESeq2 plotPCA on the 500 most variable genes after applying variance stabilizing transformation. Volcano plots were visualized using EnhancedVolcano (v1.14.0) and area-proportional Euler diagrams were made using eulerr (v6.1.1). Differential abundance analysis results are freely available at Zenodo (doi:10.5281/zenodo.7953707).

### Gene set enrichment and overrepresentation

Fgsea^30^ (v1.22.0) was used for preranked gene set enrichment analysis based on log_2_ transformed fold changes between cancer and controls obtained from DESeq2 differential abundance analysis. Significant enrichment was defined by a false discovery rate ≤ 0.05. Hallmark^9^ and Canonical Pathways gene sets derived from Kyoto Encyclopedia of Genes and Genomes (KEGG)^10^, http://www.pathway.jp, were obtained via the Molecular Signatures Database, MSigDB (v2022.1)^31^. Besides these MSigDB gene sets, custom tissue and cell type specific gene sets were made using inhouse and external references as described below. Gene set enrichment analysis results and custom gene sets are available at Zenodo (doi:10.5281/zenodo.7953707). Functional analysis of mRNA sets was done using the functional annotation tool of DAVID (v2021)^32,33^ and - for custom gene sets - the fora function of fgsea. For each functional analysis, a background was provided which included all mRNAs satisfying the count threshold in at least one sample.

#### RNA Atlas cell type gene sets

These cell type gene sets were defined based on total RNA sequencing data of cells and cell lines in the RNA Atlas^34^. Protein coding gene inclusion criteria were an expression of at least 16 counts after variance stabilizing transformation and an expression fold change of at least 10 compared to the cell type with second highest expression. 32 different cell type sets were defined, of which 15 had more than 5 genes, yet the largest gene set only included 10 genes.

#### Tabula Sapiens cell type gene sets

These cell type gene sets were based on the Tabula Sapiens v1 basis matrix defined by Vorperian et al^2^. Protein coding genes with at least 5 CPM and an expression fold change of at least 10 compared to the cell type with the second highest expression were retained. In total, 39 gene sets were obtained, of which 13 sets had more than 5 genes. The largest gene set contained 26 genes.

#### GTEX tissue gene sets

These tissue gene sets were defined using SPECS specificity scores^35^ of GTEx v7^36^ tissue samples, summarized in https://specs.cmgg.be. Criteria for protein coding gene inclusion in a certain tissue gene set were a SPECS score ≥ 0.98 and a median TPM ≥ 5. Large gene sets were reduced to the top 200 genes based on the highest SPECS score. 23 out of 27 defined tissues gene sets included more than 5 genes and the median gene set size was 50.

#### HPA tissue gene sets

These tissue gene sets were defined based on expression data from the Human Protein Atlas^37^ (HPA, v23, https://v23.proteinatlas.org/). Human Protein Atlas protein coding genes needed to have a TPM value of at least 5 and expression that is at least 10 times higher in the tissue with highest expression compared to the tissue with second highest expression. Finally, large gene sets were reduced to the top 200 genes based on the highest fold changes. Gene sets for 30 tissues were defined, with 21 gene sets including more than 5 genes and a median gene set size of 8.

#### TCGA cancer tissue gene sets

These gene sets were defined similarly to the GTEX gene sets but using expression in The Cancer Genome Atlas (TCGA) cancer tissue samples, generated by the TCGA Research Network: https://www.cancer.gov/tcga. Protein coding genes with a SPECS score of at least 0.98 in a certain tissue were included in the gene set with a limit of 200 genes, based on highest SPECS score. 23 cancer tissue gene sets were obtained with a median gene set size of 21 genes. 19 out of 23 gene sets included more than 5 genes.

### Cell type deconvolution

NuSVR deconvolution using the Tabula Sapiens v1 basis matrix was performed as described by Vorperian et al.^2^ and the accompanying GitHub repository, https://github.com/sevahn/deconvolution. As recommended by the authors, data was CPM normalized and no log transformation was performed. Note that the erythrocyte fraction is underestimated due to the absence of hemoglobin probes in mRNA capture sequencing.

### Tail genes

To identify tail genes, transcript counts of every sample were first converted to a customized z-score based on the mean and standard deviation in the reference group, including all control samples except the sample of interest. More specifically, a pseudocount was added to all normalized transcript counts followed by log_2_ transformation. Then, for each gene, the mean and standard deviation of these transformed counts in the reference group was used to scale the transformed counts in the sample of interest.

Tail genes were then defined as genes with at least 40 counts after DESeq2 normalization and deviating more than 3 standard deviations from the mean of the control reference (|z| > 3). For each gene in a sample of interest:

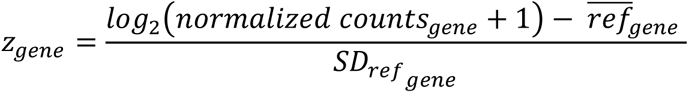

where 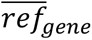 and *SD_refgene_* are respectively the mean and standard deviation of log_2_ transformed counts for this gene over all (other) control samples.

Fisher’s exact tests (based on tail gene characteristics (|z| > 3 and normalized counts ≥ 40) and donor cancer/control status) were used to select a subset of tail genes, named biomarker tail genes (BTG), that are specifically associated to the disease state.

Binary logistic regression, R glm function with binomial family, was used to classify a sample as cancer or control based on the number of biomarker tail genes. 10x 5-fold cross-validation was used: each time 80% of data (stratified partitioning) was used for BTG identification and model training, the remaining 20% was used to test the classification performance with that particular BTG set. Note that the reference used for z-score calculation never included controls from the test set. For sex-specific cancer types, only samples from control donors with the same sex were used for training and testing. Based on these predictions, the Receiver Operating Characteristic (ROC) curve and Area Under Curve (AUC) were calculated and the mean was visualized using pROC (v1.18.0)^38^.

Cross-validation resulted in different BTG sets. Consensus BTG were defined as BTG identified in at least half of the cross-validation iterations (≥25 out of 50). The number of tail genes per sample that belong to this consensus set was visualized, where the z-scores were again based on the reference with all controls from the same cohort, except the sample itself.

For internal verification, all sample labels were randomly reshuffled using the sample function in R without replacement. Next, tail gene identification and evaluation were done based on the new class labels (using the new control samples as reference). This process was repeated 20 times. For 10 iterations of label swapping, we also did biomarker tail gene identification and evaluation with 10x 5-fold cross-validation.

### Statistics

Kruskal-Wallis tests were used to compare multiple groups, and Wilcoxon rank-sum tests were used to compare two groups. Corresponding statistical significance, effect sizes and confidence intervals were calculated with rstatix (v0.7.2). For individual testing, p-values smaller than 0.05 were considered significant. In case of multiple testing, the Benjamini-Hochberg procedure was used to calculate false discovery rate adjusted p-values (q-values) and significance was defined as q smaller than 0.05.

Significance of overlap between gene sets was determined by Fisher’s exact test in GeneOverlap v1.32.0. The universe consisted of all genes considered for the corresponding differential abundance or tail gene analyses (see filtering above). A Jaccard index was calculated to assess similarity between the gene sets, with 0 indicating no similarity and 1 indicating identical sets. Jonckheere-Terpstra test was used to assess whether there was a significant trend between the cancer stage and the number of biomarker tail genes in cancer samples.

## Supporting information

Supplementary Info (including Suppl.Table 2&8)

Supplementary Analysis 1

Supplementary Analysis 2

Supplementary Analysis 3

## Data availability

Raw RNA-sequencing data is available under restricted access in the European Genome-phenome Archive (EGA) to preserve individuals’ privacy and comply with patient consent for data sharing under the European General Data Protection Regulation. The pan-cancer and three-cancer cohort FASTQs can be found under study EGAS00001006755 (dataset EGAD00001009713). The lymphoma cohort FASTQs can be found under study EGAS00001007127 (dataset EGAD00001010259). The bladder cancer cohort FASTQs can be found under study EGAS00001003917 (dataset EGAD00001005439). Differential abundance and gene set enrichment analysis results are available (open access) in a Zenodo repository (doi:10.5281/zenodo.7953707). An overview of tail genes per type and 50% consensus biomarker tail genes can be found in Supplementary Table 7.

## Code availability

Scripts used for differential abundance and tail gene analyses are available on GitHub (https://github.com/OncoRNALab/tailgenes).

## Acknowledgements

Figures 1a and 5b were created with <u>BioRender.com</u>. We would like to thank Tim Mercer for providing the Sequin spikes, Eva Hulstaert and Francisco Avila Cobos for their support during the first phase of the project, and Anneleen Decock for her feedback on the initial draft. We would also like to thank the Biostatistics Unit of the Faculty of Medicine and Health Sciences (Ghent University) for their feedback on the tail gene performance assessment.

## Author contributions

Conceptualization: AM, CE, JV, PM

Data Curation: AM

Formal analysis: AM, OT

Funding acquisition: AM, JV, JVD, PM

Investigation: EV, JN, KV

Methodology: AM, JV, PM

Project administration: AM, JV, PM

Resources: FO, JV, JVD, KS, PD, PM

Software: AM, JA Supervision: JV, PM Validation: AM, OT Visualization: AM

Writing – original draft: AM, JV, PM

Writing – review & editing: all authors

