## Extended Data Figures for "Patient-specific alterations in blood plasma cfRNA profiles enable accurate classification of cancer patients and controls"

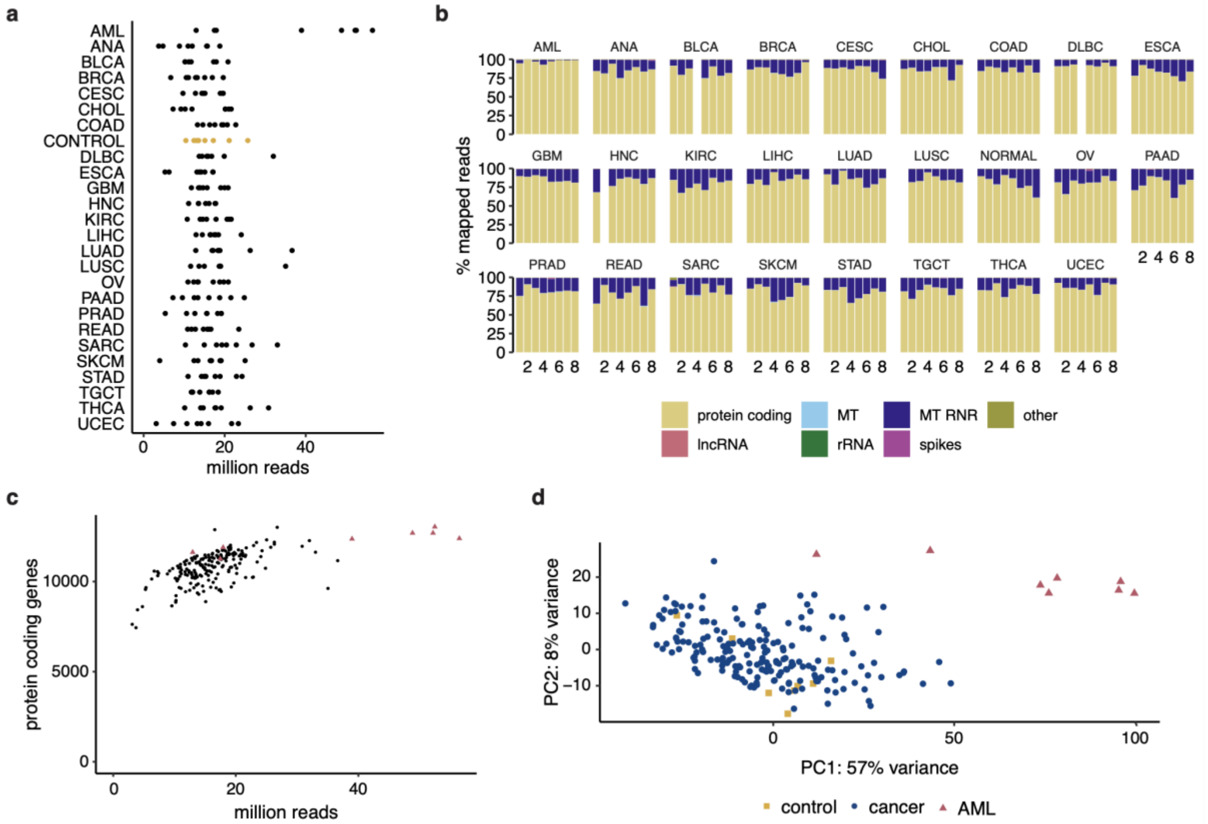


***Extended Data Fig.1: MRNA capture sequencing data characteristics pan-cancer cohort. a,*** *Sequencing depth. Plot shows million paired-end reads per sample after quality filtering. Groups ordered alphabetically. No significant difference between groups: Kruskal-Wallis test p = 0.0503, with moderate effect size (0.071), smallest post-hoc two-sided Wilcoxon rank-sum test q = 0.354.* ***b****, Capture sequencing resulted in mRNA-enriched plasma cfRNA profiles. RNA biotype distribution of mapped reads in pan-cancer cohort. Each bar represents one sample. Samples with less than 2M reads were excluded. Protein coding: protein coding gene transcripts (mRNA); lncRNA: long non-coding RNA; MT: mitochondrial RNA; MT RNR: mitochondrially encoded ribosomal RNA; rRNA: ribosomal RNA; spikes: Sequin spike-in RNA; other: other RNA transcripts.* ***c*** *and* ***d****, Deeper sequencing of AML samples does not drive results.* ***c:*** *Sequencing depth and number of protein coding genes are only moderately correlated (spearman correlation r = 0.63, p < 2.2E-16). Red triangles: acute myeloid leukemia samples; black dots: non-leukemia cancer and control samples. Median number of protein coding genes above detection threshold in AML samples is 12,342 and 11,239, respectively, without and with downsampling reads to 16M (median reads per sample in cohort).* ***d****: Similar principal component analysis results as in Fig.1d when downsampling AML sample reads to median reads in cohort (16M). 500 most variable mRNAs in the pan-cancer cohort used (with variance stabilizing transformation DESeq2).*

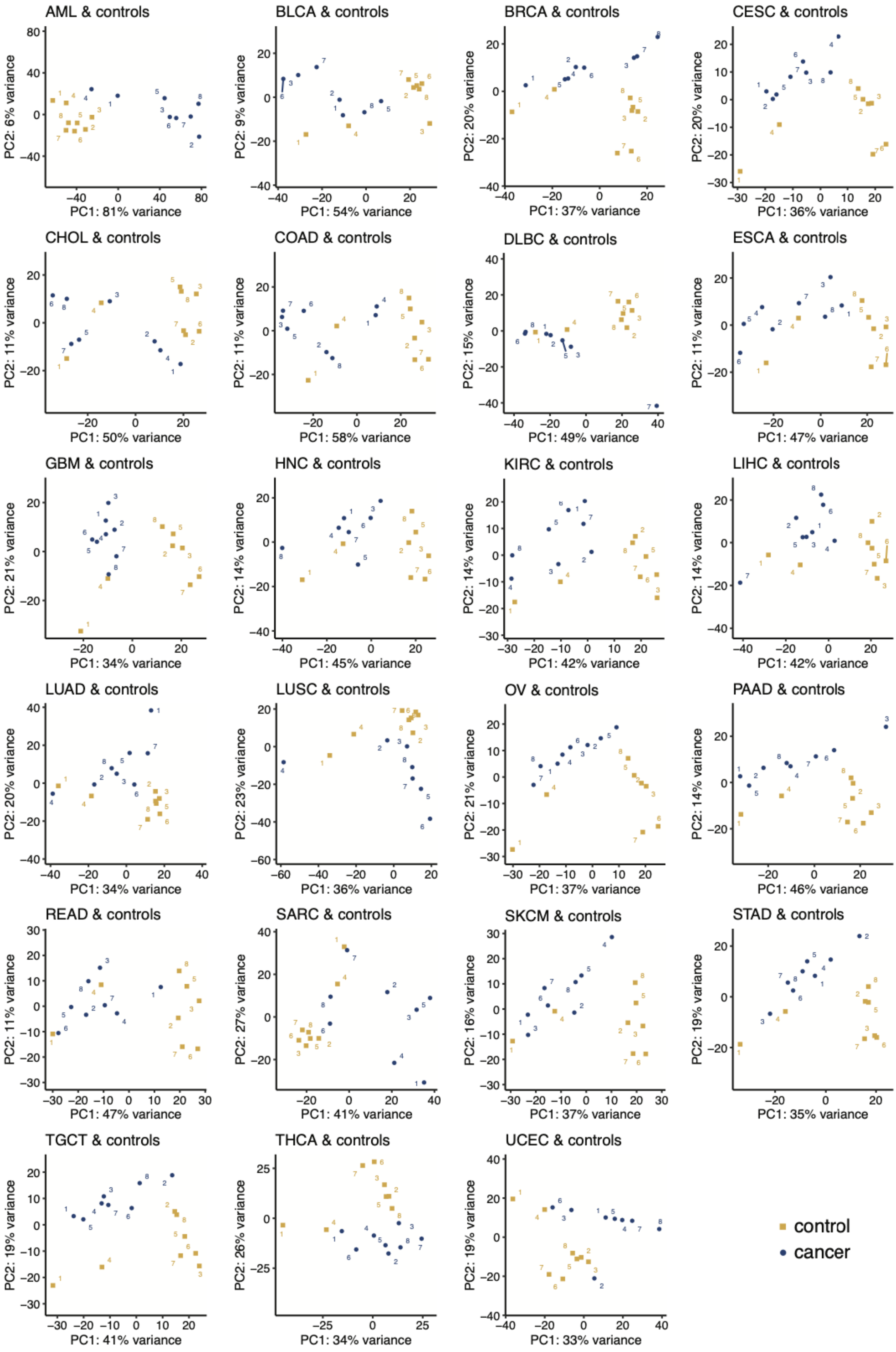
AML & controls
PC1: 81% variance
PC2: 6% variance
BLCA & controls
PC1: 54% variance
PC2: 9% variance
BRCA & controls
PC1: 37% variance
PC2: 20% variance
CESC & controls
PC1: 36% variance
PC2: 20% variance
CHOL & controls
PC1: 50% variance
PC2: 11% variance
COAD & controls
PC1: 58% variance
PC2: 11% variance
DLBC & controls
PC1: 49% variance
PC2: 15% variance
ESCA & controls
PC1: 47% variance
PC2: 11% variance
GBM & controls
PC1: 34% variance
PC2: 21% variance
HNC & controls
PC1: 45% variance
PC2: 14% variance
KIRC & controls
PC1: 42% variance
PC2: 14% variance
LIHC & controls
PC1: 42% variance
PC2: 14% variance
LUAD & controls
PC1: 34% variance
PC2: 20% variance
LUSC & controls
PC1: 36% variance
PC2: 23% variance
OV & controls
PC1: 37% variance
PC2: 21% variance
PAAD & controls
PC1: 46% variance
PC2: 14% variance
READ & controls
PC1: 47% variance
PC2: 11% variance
SARC & controls
PC1: 41% variance
PC2: 27% variance
SKCM & controls
PC1: 37% variance
PC2: 16% variance
STAD & controls
PC1: 35% variance
PC2: 19% variance
TGCT & controls
PC1: 41% variance
PC2: 19% variance
THCA & controls
PC1: 34% variance
PC2: 26% variance
UCEC & controls
PC1: 33% variance
PC2: 19% variance
control
cancer

***Extended Data Fig.2: Most cancer and control samples are separated in the first two principal components when considering individual types.*** *Principal component analysis using the 500 most variable mRNAs based on samples of control group and one cancer type (with variance stabilizing transformation DESeq2). Replicate numbers are indicated in the plot. Yellow squares: control samples; blue dots: cancer samples.*

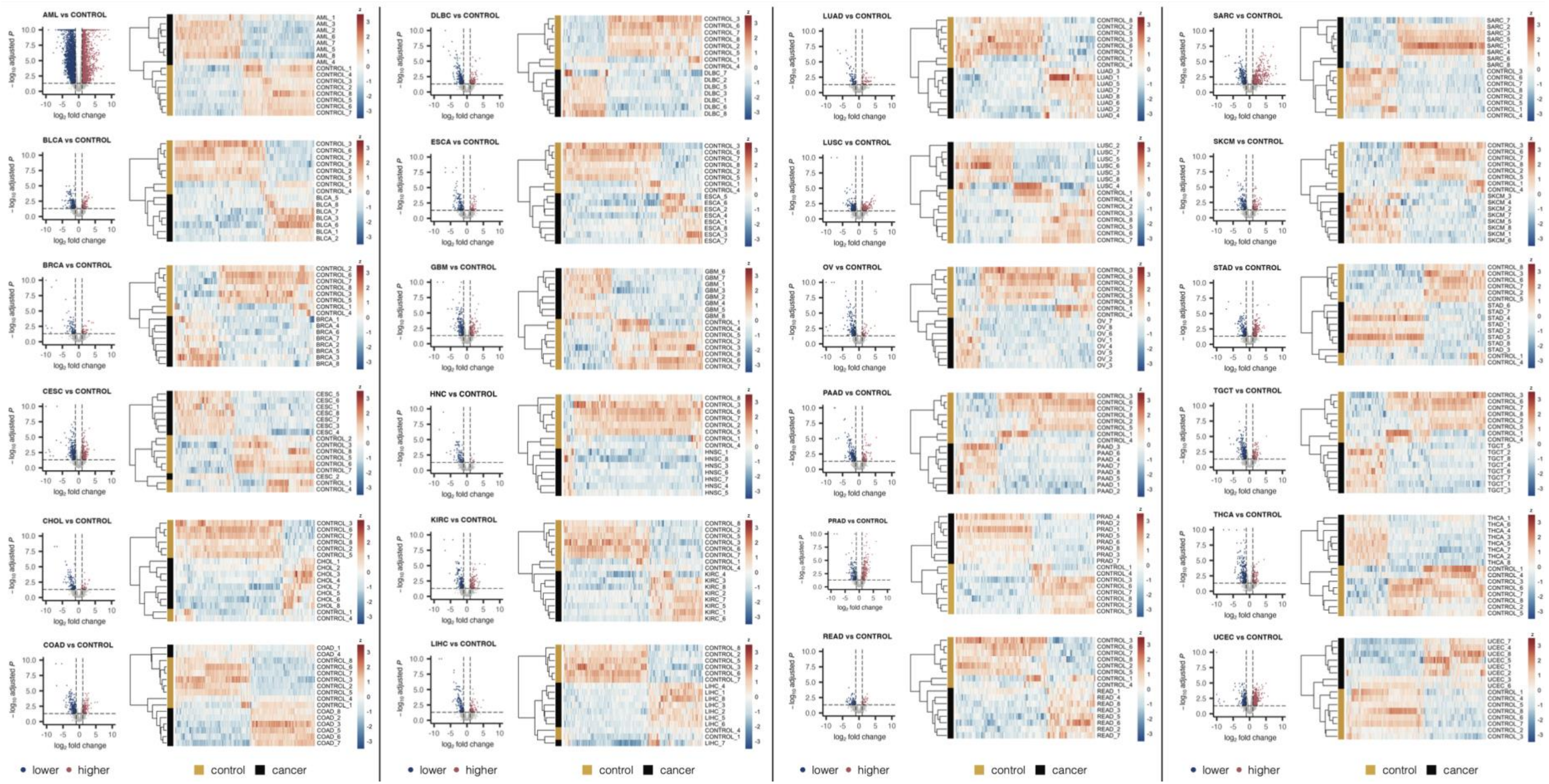


***Extended Data Fig.3: Differentially abundant mRNAs in plasma of cancer patients compared to controls.*** *Volcano plots show fold change and multiple testing corrected p-values of mRNAs in cancer versus control. Horizontal line at adjusted p-value of 0.05 and vertical lines at log2 fold change of -1 and 1, resp. Higher (red): higher abundant in cancer compared to control samples (q < 0.05 and $log_2$ fold change > 1); lower (blue): lower abundant in cancer compared to control samples (q < 0.05 and $log_2$ fold change < -1). Heatmaps show relative abundance of differentially abundant genes (columns) in individual plasma samples (rows) based on log2(normalized counts +1) followed by z-score transformation per gene. Clustering of genes and samples based on Pearson correlation.*

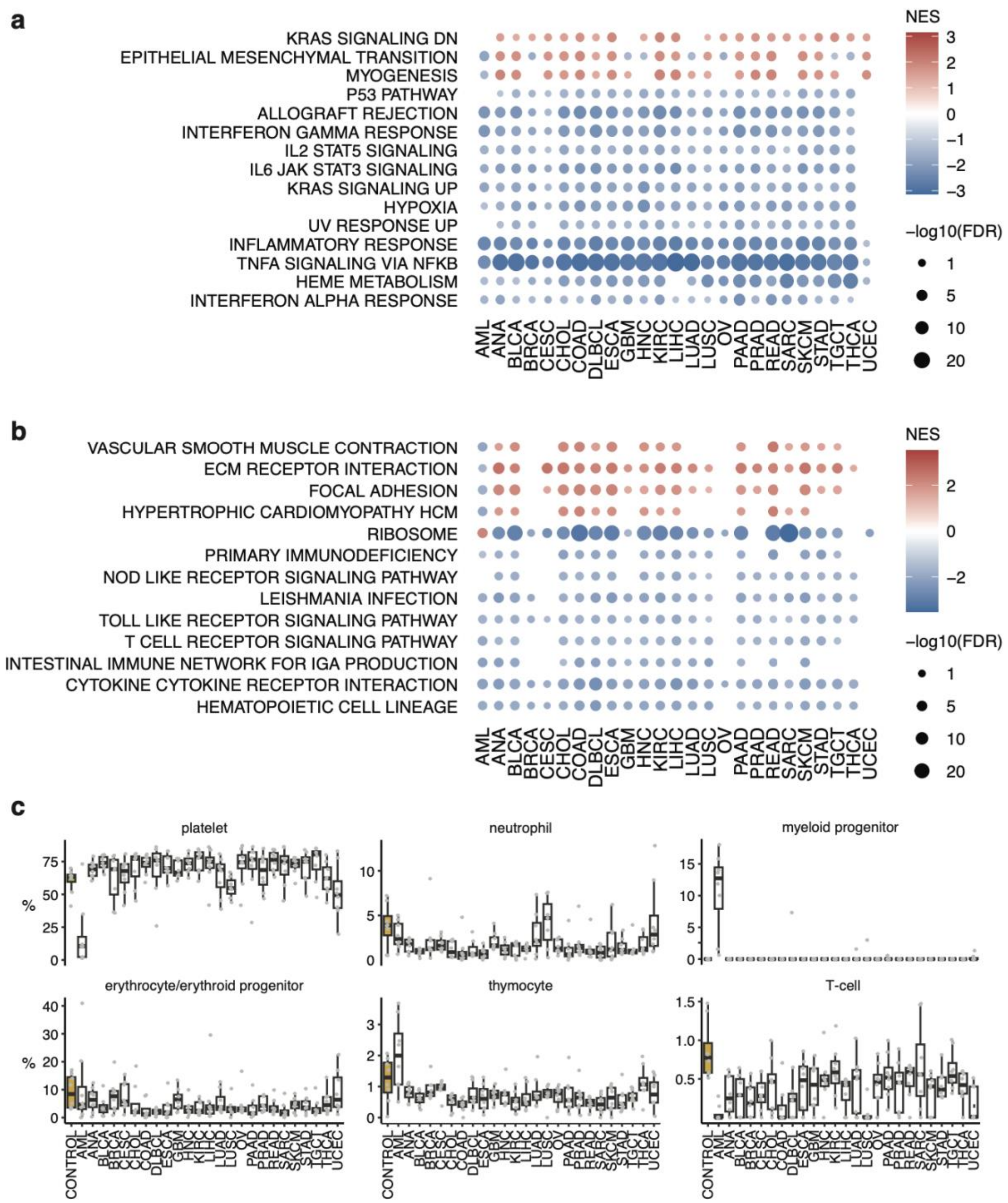


***Extended Data Fig.4: Recurrent patterns in gene set enrichment and deconvolution analyses. a** and **b**, gene set enrichment analysis for hallmark gene sets (**a**) and KEGG canonical pathway genes (**b**). Only pathways that are significantly enriched in at least half of the cancer types are shown. Colored according to normalized enrichment scores (NES) for gene sets with q < 0.05. **c,** Cell type contributions to plasma cfRNA profiles show distinct leukemic signals and lower immune cell fractions in cancer samples. Boxplots of specific cell fractions based on deconvolution. Boxplots show lower quartile (Q1), median, and upper quartile (Q3). Whiskers extend from the lower and upper quartile to the smallest and largest value, respectively, within at most 1.5*interquartile range (Q3-Q1) from that quartile. More extreme points are plotted as individual dots. Yellow: control samples; grey*

*dots: individual sample fractions. Cell type fractions obtained by nuSVR deconvolution using Tabula Sapiens v1.0 as basis matrix.*

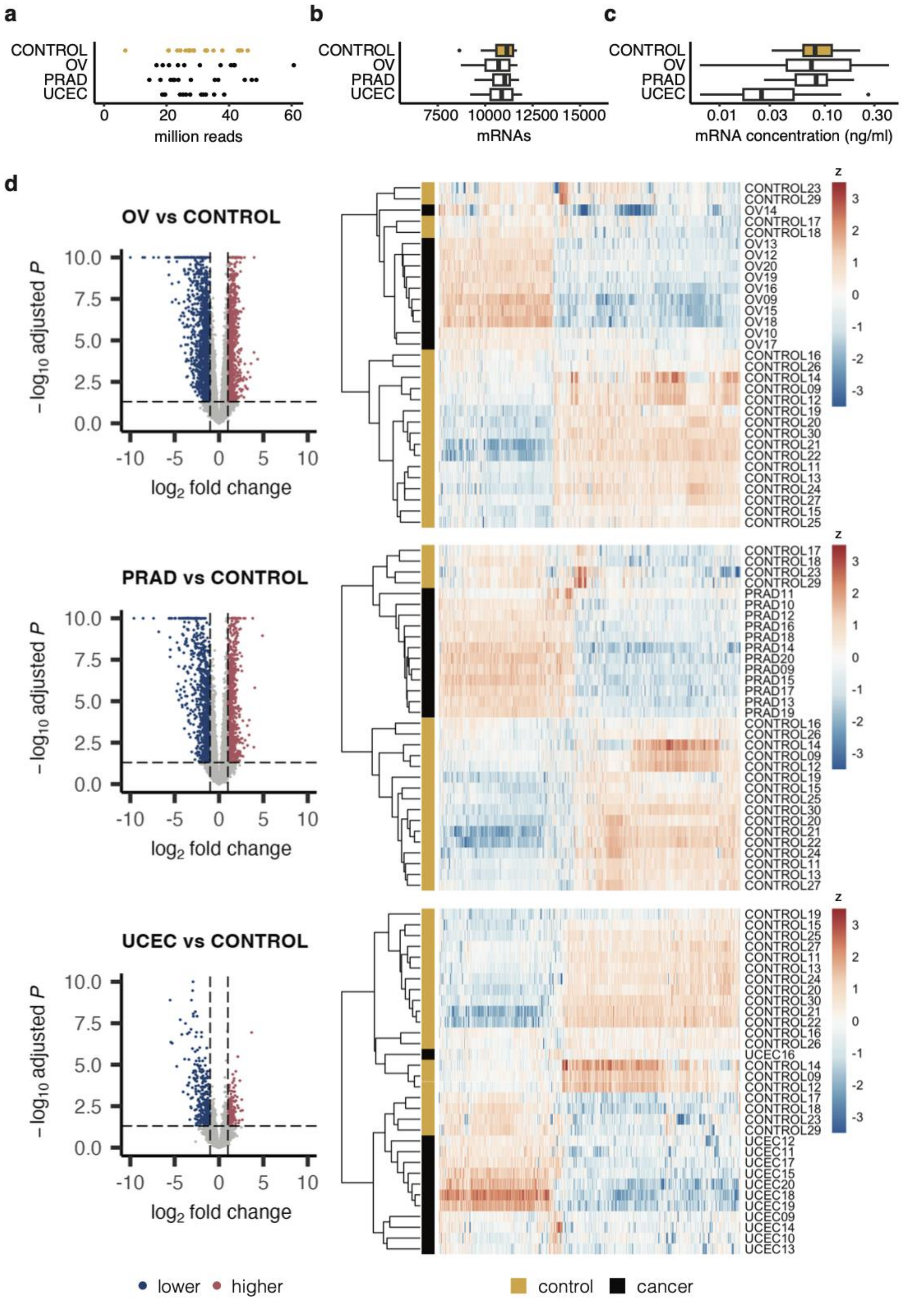


***Extended Data Fig.5: MRNA capture sequencing data characteristics and differentially abundant mRNAs in three-cancer cohort. a**, Plot shows million paired-end reads after quality filtering with no significant difference*

between groups (Kruskal-Wallis $p = 0.7858$). **b**, Number of messenger RNAs (mRNAs) with at least 10 counts, no significant difference between groups (Kruskal-Wallis $p = 0.8116$). **c**, mRNA concentration in plasma based on Sequin spikes with no significant difference between groups (Kruskal-Wallis $p = 0.05902$). **b** and **c**, Boxplots show lower quartile (Q1), median, and upper quartile (Q3). Whiskers extend from the lower and upper quartile to the smallest and largest value, respectively, within at most 1.5*interquartile range (Q3-Q1) from that quartile. More extreme points are plotted as individual dots. Groups ordered alphabetically and controls indicated in yellow. **d**, differentially abundant mRNAs in plasma of cancer patients compared to controls. Volcano plots show fold change and multiple testing corrected p-values of mRNAs in cancer versus control. Horizontal line at adjusted p-value of 0.05 and vertical lines at fold change 0.5 and 2. More (red): more abundant in cancer compared to control samples ($q < 0.05$ and fold change $> 2$); less (blue): less abundant in cancer compared to control samples ($q < 0.05$ and fold change $< 0.5$). Heatmaps show relative abundance of differentially abundant genes (columns) in individual plasma samples (rows) based on log2(normalized counts +1) followed by z-score transformation per gene. Clustering of genes and samples based on Pearson correlation.

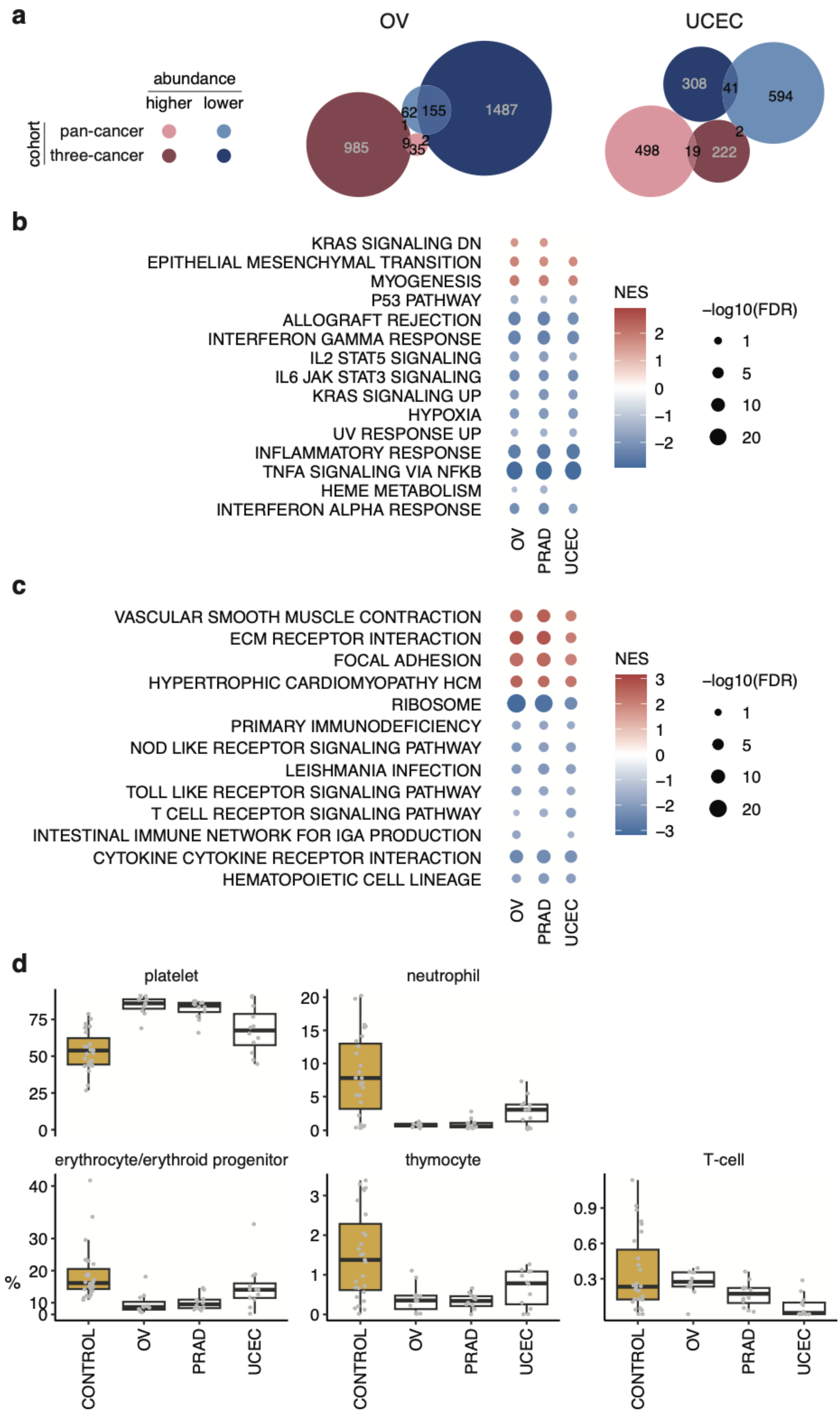

a
OV
UCEC
abundance
higher
lower
cohort
pan-cancer
three-cancer
985
62
155
1487
1
9
2
35
308
41
594
498
19
222
2
b
KRAS SIGNALING DN
EPITHELIAL MESENCHYMAL TRANSITION
MYOGENESIS
P53 PATHWAY
ALLOGRAFT REJECTION
INTERFERON GAMMA RESPONSE
IL2 STAT5 SIGNALING
IL6 JAK STAT3 SIGNALING
KRAS SIGNALING UP
HYPOXIA
UV RESPONSE UP
INFLAMMATORY RESPONSE
TNFA SIGNALING VIA NFKB
HEME METABOLISM
INTERFERON ALPHA RESPONSE
OV
PRAD
UCEC
NES
–log10(FDR)
c
VASCULAR SMOOTH MUSCLE CONTRACTION
ECM RECEPTOR INTERACTION
FOCAL ADHESION
HYPERTROPHIC CARDIOMYOPATHY HCM
RIBOSOME
PRIMARY IMMUNODEFICIENCY
NOD LIKE RECEPTOR SIGNALING PATHWAY
LEISHMANIA INFECTION
TOLL LIKE RECEPTOR SIGNALING PATHWAY
T CELL RECEPTOR SIGNALING PATHWAY
INTESTINAL IMMUNE NETWORK FOR IGA PRODUCTION
CYTOKINE CYTOKINE RECEPTOR INTERACTION
HEMATOPOIETIC CELL LINEAGE
OV
PRAD
UCEC
NES
–log10(FDR)
d
platelet
neutrophil
erythrocyte/erythroid progenitor
thymocyte
T-cell
%
CONTROL
OV
PRAD
UCEC

***Extended Data Fig.6: Significant, but small, overlap in differentially abundant genes between cohorts with recurrence of enrichment patterns and cell type contributions. a,*** *Overlap between differentially abundant genes in cancer versus control between pan-cancer and three-cancer cohort of ovarian (OV) and uterine cancer (UCEC), respectively. Fisher's exact test OV: p = 4.0E-4, odds ratio = 4.6, Jaccard index = 0.009 for higher abundant genes (higher); p = 8.4E-118, odds ratio = 30.0, Jaccard index = 0.091 for lower abundant genes (lower). Fisher's exact test UCEC: p = 2.2E-5, odds ratio = 3.3, Jaccard index = 0.026 for higher abundant genes (higher); p = 7.6E-13, odds ratio = 4.2, Jaccard index = 0.043 for lower abundant genes (lower). Circles are proportional to the number of genes within one cancer vs control comparison. Differential abundance: q < 0.05 &* $|log_2$ *fold change| > 1.* ***b*** *and* ***c****, Gene set enrichment in three-cancer cohort based on hallmark (b) and KEGG gene sets (c), respectively, obtained from Human Molecular Signatures Database***.*** *Only gene sets significantly enriched in at least half of the cancer types of the pan-cancer cohort (cf. Extended Data Fig.4) are shown. Coloring according to normalized enrichment scores (NES) for gene sets with q < 0.05.* ***d****, Cell type contributions to plasma cfRNA show increased platelet and lower immune cell fractions in cancer samples compared to controls. Boxplots of specific cell fractions obtained from nuSVR deconvolution using Tabula Sapiens v1.0 as basis matrix. Boxplots show lower quartile (Q1), median, and upper quartile (Q3). Whiskers extend from the lower and upper quartile to the smallest and largest value, respectively, within at most 1.5*interquartile range (Q3-Q1) from that quartile. More extreme points are plotted as individual dots. Yellow: control samples; grey dots: individual sample fractions.*

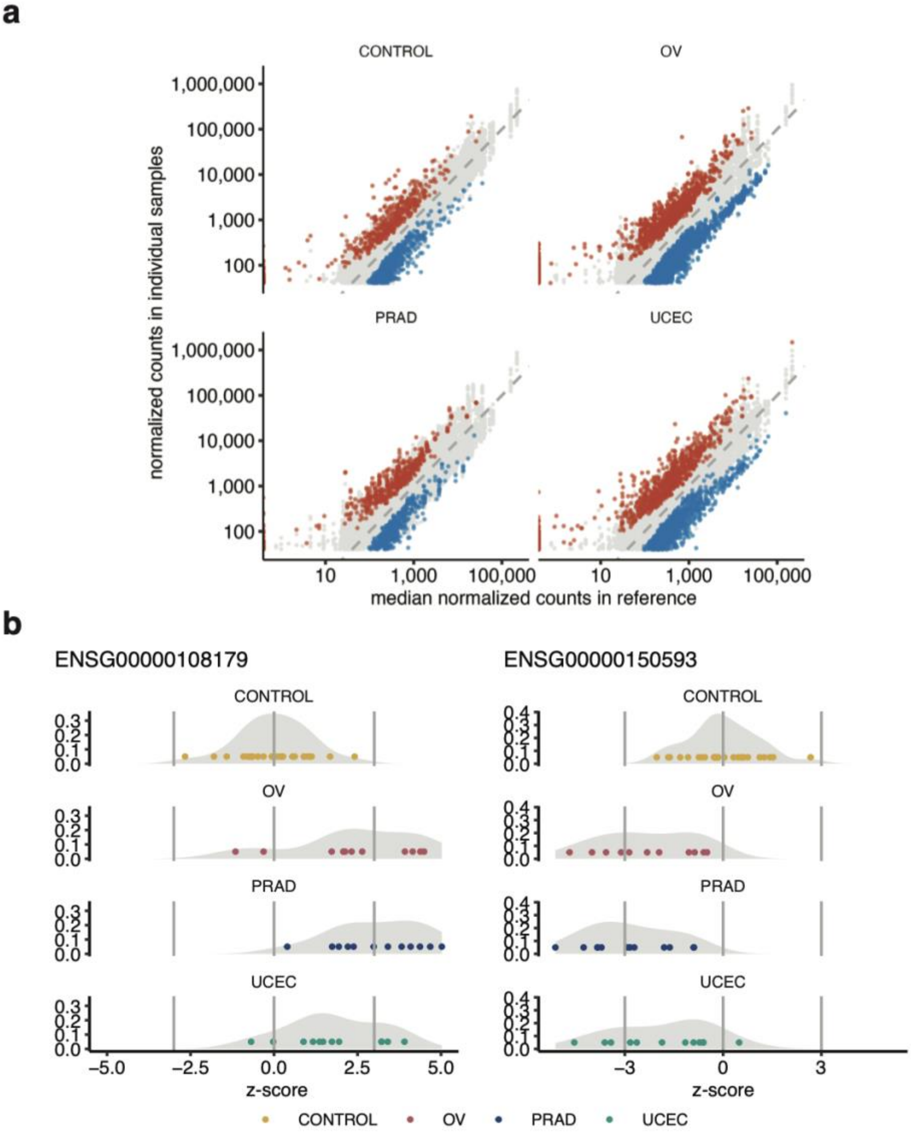


***Extended Data Fig.7: Tail gene counts and z-score distribution per type. a, for all unique tail genes:*** *normalized tail gene counts (≥ 40) in individual samples of a certain type compared to the median of the control reference (x-axis). Red (colored dots above diagonal): z > 3; blue (colored dots below diagonal): z < -3.* ***b, for two tail genes (PPIF and PDCD4) detected in multiple cancer samples:*** *z-scores per type where individual dots represent the exact z-scores for that gene in a particular sample (compared to control reference) and a grey density plot. The genes are identified as tail gene in samples where the respective z-score is smaller than -3 (left vertical line) or above 3 (right vertical line).*

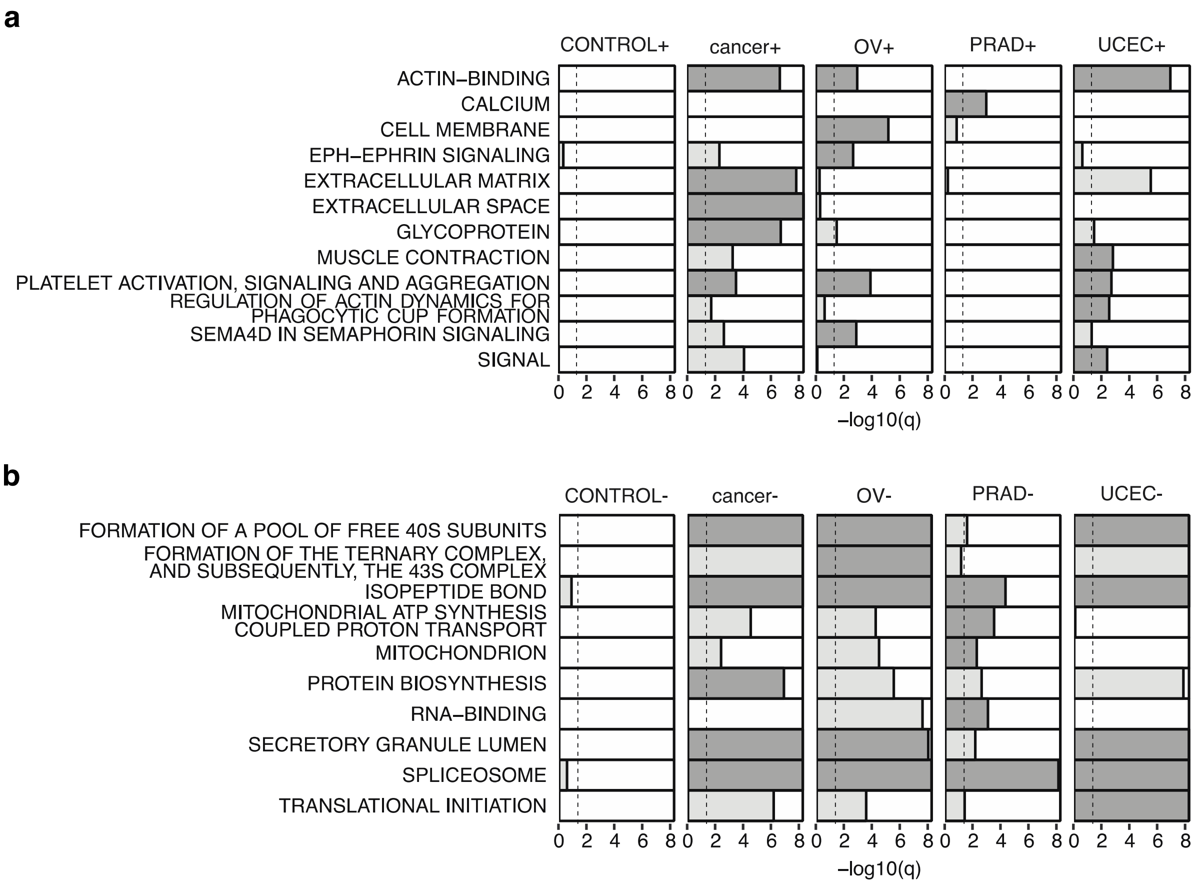


***Extended Data Fig.8: Recurrence of top overrepresented patterns in cancer sample tail genes and absence in control sample tail genes. a*** *and* ***b,*** *top 5 most enriched clusters ($q < 0.05$) for positive ($z > 3$) and negative ($z < -3$) tail genes, respectively. Dark grey: -$\log_{10}$(q-value) of the most significant annotation (Benjamini correction) for each of the 5 most enriched clusters per group of tail genes (CONTROL, OV, PRAD, UCEC, or 'cancer', i.e. all three cancer types combined) based on DAVID functional clustering analysis. Light grey: -log10(q-value) of top-5 annotations of other groups that are not in top-5 of the OV, PRAD, UCEC, all cancer, or CONTROL group. -log10(q) topped of at 8. Dashed vertical lines at $q = 0.05$.*

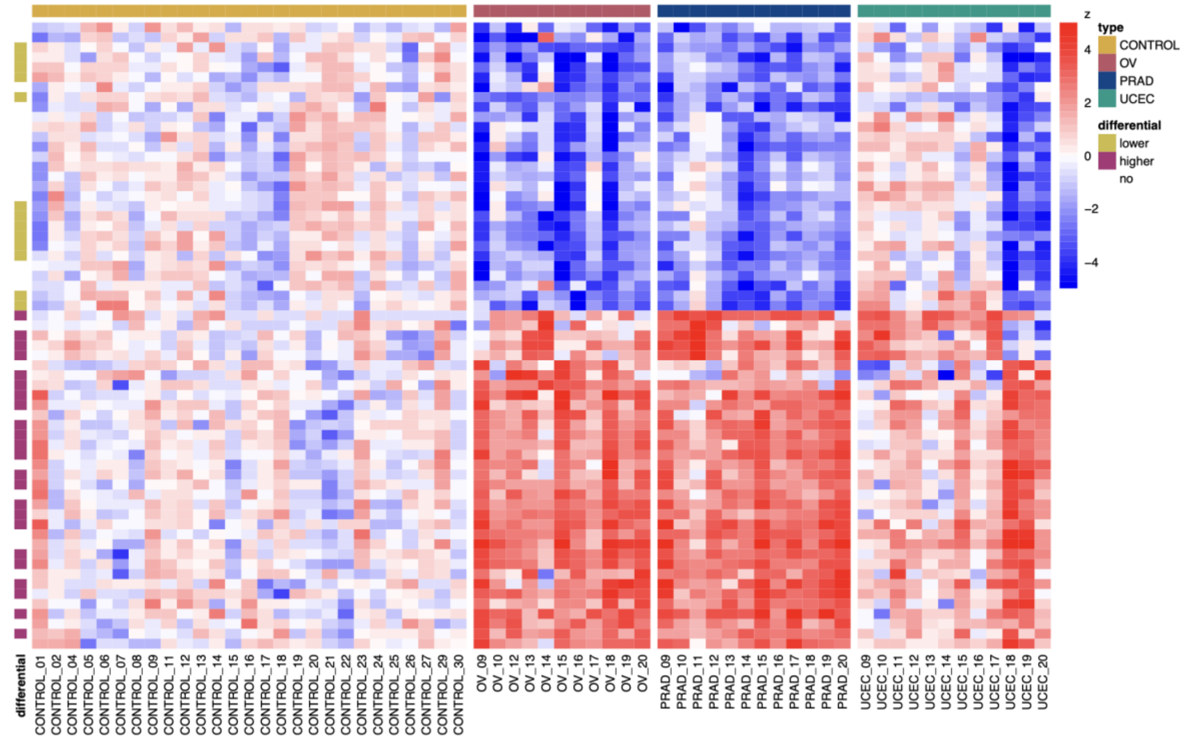


***Extended Data Fig.9: Z-score distribution of selected tail genes in the three-cancer cohort.*** *Heatmap shows z-scores of the 63 consensus tail genes, genes identified as biomarker tail genes in at least half of the iterations (10x 5-fold cross-validation). Genes (rows) clustered based on Euclidean distance.*

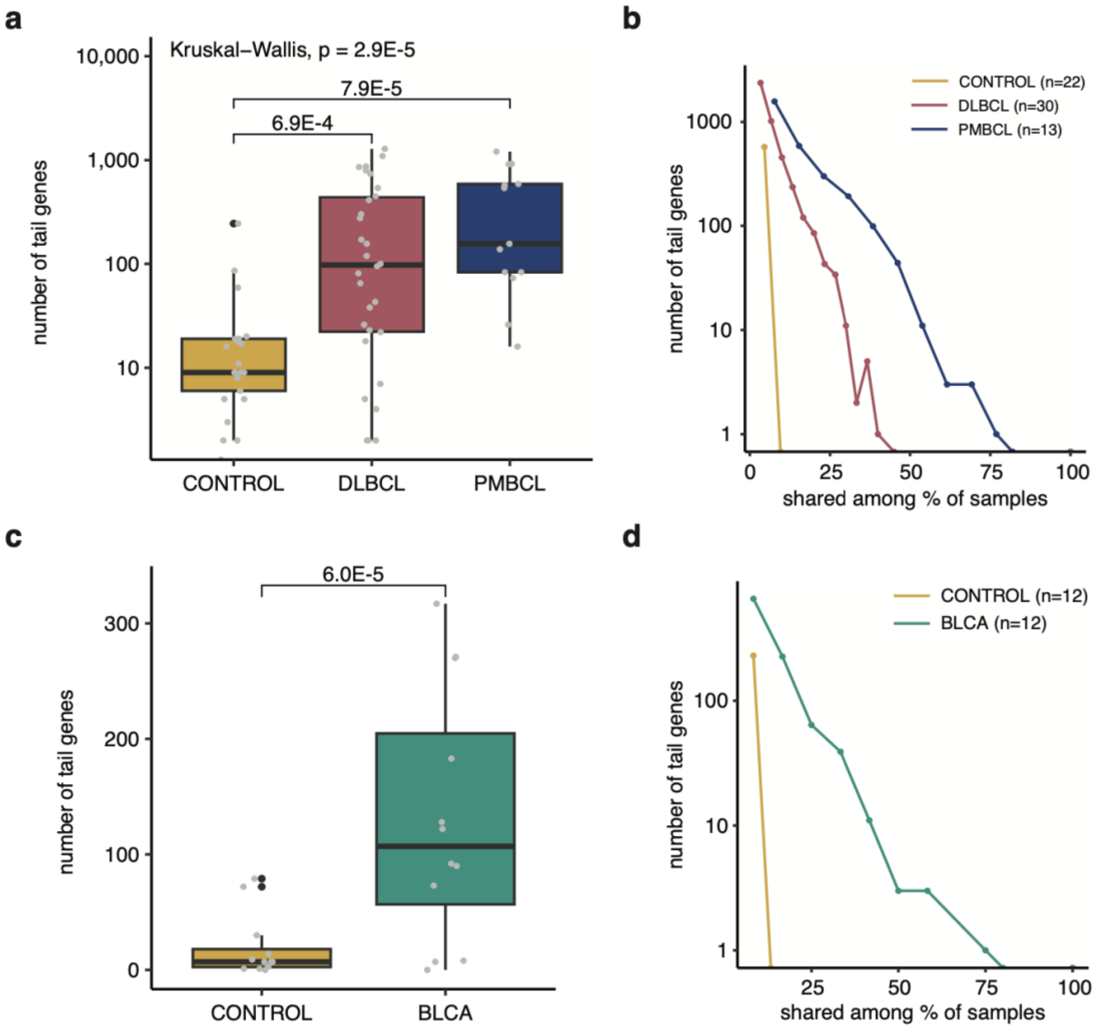


***Extended Data Fig.10: Tail genes in the lymphoma plasma and bladder cancer urine cohort. a** and **b**, lymphoma plasma cohort: plasma samples from 22 control (CONTROL), 30 diffuse large B-cell lymphoma (DLBCL), and 13 primary mediastinal large B-cell lymphoma (PMBCL) patients. **a**, Boxplot shows number of tail genes (n = 5374) per group (lower quartile, median, upper quartile, whiskers of 1.5*interquartile range, more extreme points indicated by black dots), grey dots represent individual sample counts. Wilcoxon rank-sum q-values for specific cancer versus control comparisons indicated in the plot. **b**, Number of tail genes shared by a certain fraction of samples of the lymphoma or control group. Number of samples indicated between brackets. **c** and **d**, urine cohort: urine samples from 12 control (CONTROL) and 12 bladder cancer (BLCA) patients. **c**, Boxplot shows number of tail genes (n = 1152) per group (lower quartile, median, upper quartile, whiskers of 1.5*interquartile range, more extreme points indicated by black dots), grey dots represent individual sample counts. **d**, Number of tail genes shared by a certain fraction of samples of the bladder cancer or control group. Number of samples indicated between brackets.*
