## Supplementary Info (including Suppl.Table 2&8) for "Patient-specific alterations in blood plasma cfRNA profiles enable accurate classification of cancer patients and controls"

### Supplementary Information

#### Supplementary Tables

***Supplementary Table 1: Overview of donor, disease, and sample information***. *Cohort: sample cohort (Table1) to which a certain plasma sample belongs; DonorID: donor code; RNAID: sample code; Cancer: cancer type; Abbreviation: cancer type abbreviation; ReplicateNr: replicate number; CollectionDate: date of sample collection (blood draw and plasma preparation); CollectionTube: blood collection tube; Biofluid: biofluid type of sample; Age, Sex, Ethnicity: age, sex (F for female, M for male), and ethnicity of the sample donor; TumorLocation; HistologicalDiagnosis; Grade; TNM; Stage; Treatment: ongoing treatment; Excluded: indicates samples excluded based on insufficient sequencing reads (< 2M), see Methods.*

***Supplementary Table 2: Fusion genes of TCGA found in individual plasma samples.*** *Fusion: fusion gene; TCGA: cancer type(s) for which fusion gene was detected in The Cancer Genome Atlas (TCGA); PlasmaID: plasma sample id (cancer type followed by replicate number) for which the fusion transcript is detected by FusionCatcher; SpanningPairs: Count of pairs of reads supporting the fusion; SpanningUniqueReads: Count of unique reads mapping on the fusion junction.*

| Fusion | TCGA | PlasmaID | SpanningPairs | SpanningUniqueReads |
|---|---|---|---|---|
| *PML::RARA* | AML | AML_5 | 19 | 6 |
| *RARA::PML* | AML | AML_5 | 19 | 17 |
| *PML::RARA* | AML | AML_8 | 7 | 9 |
| *SOS1::MAP4K3* | TGCT, THCA | CESC_2 | 1 | 3 |
| *SOS1::MAP4K3* | TGCT, THCA | CESC_6 | 2 | 5 |
| *PPA2::TBCK* | LUAD | STAD_2 | 1 | 4 |
| *SPAG9::MBTD1* | OV, STAD | LIHC_2 | 1 | 2 |
| *LTBP1::BIRC6* | BRCA, AML, LUSC, PRAD | LIHC_7 | 1 | 2 |
| *TAB3::DMD* | SARC | KIRC_1 | 1 | 3 |
| *MITF::FOXP1* | SKCM | KIRC_4 | 1 | 2 |
| *TNRC6A::PRKCB* | GBM | LUAD_7 | 1 | 3 |
| *CLEC16A::TXNDC11* | GBM | PAAD_2 | 1 | 3 |
| *LTBP1::BIRC6* | BRCA, AML, LUSC, PRAD | READ_8 | 1 | 3 |
| *ASAP2::MBOAT2* | LUAD | SARC_4 | 2 | 3 |
| *SNX6::BAZ1A* | STAD | SARC_6 | 1 | 2 |
| *DENND3::PTK2* | GBM | TGCT_7 | 1 | 2 |
| *CTBP2::MGMT* | LGG | PRAD_18 | 1 | 3 |

***Supplementary Table 3: Overrepresentation of Reactome, KEGG and Gene Ontology gene sets in recurrent lower abundant mRNAs.*** *DAVID functional annotation chart for the (26) mRNAs with q < 0.05 and log2 fold change < -1 in cancer versus control for at least 23 out of 25 cancer types of the pan-cancer cohort. Category: original database/resource where the term orients, Term: enriched term associated with input gene list, Count: number of input genes involved in the term, Percentage: involved genes divided by total input genes, P-value: EASE Score – modified Fisher Exact p-value, Genes: involved gene names, List total: number of input genes that are annotated to at least one term in the category. Pop hits: number of background genes annotated to the specific term. Pop total: number of background genes annotated to at least one term in the category, Fold Enrichment, Bonferroni, Benjamini, FDR (false discovery rate).*

***Supplementary Table 4: Cell type deconvolution results.*** *Fractions obtained by nuSVR deconvolution using Tabula Sapiens v1.0 as basis matrix. Pan-cancer: deconvolution results for the plasma samples of the pan-cancer cohort. Three-cancer: deconvolution results for the independent plasma samples of the three-cancer cohort.*

***Supplementary Table 5: DAVID functional annotation clustering based on tail gene lists.*** *Anycancer+, OV+, PRAD+, UCEC+, CONTROL+: based on tail genes with z > 3 in any cancer, OV, PRAD, UCEC, CONTROL sample of the three-cancer cohort, respectively. Anycancer-, OV-, PRAD-, UCEC-, CONTROL-: based on tail genes with z < -3 in any cancer, OV, PRAD, UCEC, CONTROL sample of the three-cancer cohort, respectively. Category: original database/resource where the term orients, Term: enriched term associated with input gene list, Count: number of input genes involved in the term, Percentage: involved genes divided by total input genes, P-value: EASE Score – modified Fisher Exact p-value, Genes: involved gene names, List total: number of input genes that are annotated to at least one term in the category. Pop hits: number of background genes annotated to the specific term. Pop total: number of background genes annotated to at least one term in the category, Fold Enrichment, Bonferroni, Benjamini, FDR (false discovery rate).*

***Supplementary Table 6: Overview of biomarker tail gene set sizes and classification performance in the different cohorts***. *Cohort: sample cohort (Table 1). Comparison: cancer types of interest versus control samples. BTG set: biomarker tail gene set identified in training set for the respective comparison (10x 5-fold cross-validation). Mean AUC: mean of obtained AUCs for test sets. Ci: 95% confidence interval. "male" or "female": only looking at performance in test sets for samples from male or female donors, respectively.*

***Supplementary Table 7: (biomarker) tail genes***. *Cohort: sample cohort (Table 1). Type & nr_samples: cancer or control group and number of samples in this group for which the gene (in columns "ensemble gene id" & "gene name") was identified as tail gene (|z| > 3 and normalized counts ≥ 40). Pos_z & neg_z: number of samples with positive or negative z-value for tail gene. Consensus_BTG_cancer: TRUE if the gene is a 50% consensus biomarker tail gene in 1 cancer group vs control (in rows where "type" is a cancer: for that particular cancer versus control comparison; in rows where "type" is control: for any of the cancer-control comparisons in cohort).*

*Consensus_BTG_combined: TRUE if the gene is a consensus biomarker tail gene in all cancers in the cohort combined vs control.*

***Supplementary Table 8: Splicing and exon coverage***. *% spliced: percentage of total reads mapping to spliced junctions; % exons: percentage of reads mapping to exonic regions (not intronic or intergenic); sd: standard deviation.*

| **cohort** | **library preparation** | **mean % spliced** | **sd % spliced** | **mean % exons** | **sd % exons** |
|---|---|---|---|---|---|
| pan-cancer | mRNA capture | 14.78 | 3.38 | 79.82 | 3.40 |
| three-cancer | mRNA capture | 10.25 | 2.18 | 80.58 | 3.32 |
| lymphoma | total RNA | 3.88 | 1.76 | 70.60 | 12.86 |
| bladder cancer | mRNA capture | 35.83 | 2.91 | 94.92 | 4.16 |

#### Supplementary Analyses

***Supplementary Analysis 1****: Heterogeneity of cohorts based on differential abundance results.*

***Supplementary Analysis 2:*** *Biomarker tail genes linked to cancer in literature.*

***Supplementary Analysis 3****: Spike analyses.*
