## Supplementary Analysis 1 for "Patient-specific alterations in blood plasma cfRNA profiles enable accurate classification of cancer patients and controls"

### Supplementary Analysis 1 - heterogeneity

#### Introduction

In this Supplementary Analysis file we will demonstrate the effect of heterogeneity between populations/studies on the number of common discoveries. In particular, we consider the setting of two independent studies for differential gene expression (DE). Each study comes with a different sample size. We will develop a simple method for approximating the expected number of discoveries (significant DE genes) by each study separately, and the number discovered by both studies. We will develop a probabilistic model that can describe two homogeneous study populations (i.e. gene expression effect sizes are the same in the two populations), as well as two heterogeneous study populations (i.e. none of the genes have the same effect size in both populations), and everything in between.

Our calculations will demonstrate that more common discoveries are expected when the populations are similar as compared to heterogeneous populations, and that this conclusion holds for small and large sample sizes.

#### Methodology

Let $\theta_{jk}$ denote the effect size (e.g. log fold change, LFC) of gene $j = 1, \dots, p$ in study $k = 1, 2$. In study $k$ we are interested in testing the null hypotheses $H_{0jk} : \theta_{jk} = 0$, for all genes $j$.

Despite the two studies being independent, the test results are not. This can be understood as follows. Suppose that the target populations of the two studies are the same (homogeneous populations), then we have $\theta_{j1} = \theta_{j2}$. Hence, if study 1 rejects the corresponding null hypothesis (because $\theta_{j1}$ is large), then it is likely that the same null hypothesis will also be rejected in the second study, because $\theta_{j2} = \theta_{j1}$ is also large. However, if $\theta_{j1} \neq \theta_{j2}$ (population heterogeneity), and there is no relationship between the two $\theta$ parameters, then the test results of the two studies will be independent.

To allow for homegeneous and heterogeneous populations, and to allow for a simplification of the calculations, we will adopt the following setup. Assume

$$\theta_{j1} \sim F_\theta \quad \text{and} \quad \theta^*_{j2} \sim G_\theta,$$

with $F_\theta$ and $G_\theta$ two different distributions, and assume that the $\theta_{j1}$ and $\theta^*_{j2}$ are independently distributed.

Let $S_j$ denote a 0/1 binary random variable with $P(S_j = 1) = \gamma$, and define

$$\theta_{j2} = S_j\theta_{j1} + (1 - S_j)\theta^*_{j2}.$$

With this setup, the effect sizes (LFC) in the first study can be described by distribution $F_\theta$. With a probability of $\gamma$, the LFC of gene $j$ in the second study is the same as in the first, but with a probability of $1 - \gamma$ the LFC is different and given by $\theta^*_{j2}$, which behaves as $G_\theta$.

The parameter $\gamma$ reflects the degree of homogeneity: if $\gamma = 1$, then the two study populations are identical, and if $\gamma = 0$, then all genes show different effect sizes in the two populations.

In this document we assume zero-mean normal distributions for $F_\theta$ and $G_\theta$. This will facilitate the calculations.

Based on this model, we can calculate the expected number of detected DE genes in study 1 ($E_1$), in study 2 ($E_2$) and the expected number of genes that are detected by both methods ($E$). Details of the calculation are given at the end of this document (Appendix). The calculations require also other parameters as input:

variances of the gene expressions for the two studies, variances (over the genes) of the effect sizes for the two studies, the number of genes analysed in both studies, and the nominal FDR level (here: 5%). These parameters were estimated from the data.

### Results

For each of the three cancer types we show graphs showing the effect of study heterogeneity (parameter $\gamma$) on the number of DE genes discovered in both studies. In particular, the overlap between the two sets of DE genes will be quantified by the Jaccard index, which is defined as the number of common discoveries, divided by the total number of discoveries (in both studies):

$$J = \frac{E}{E_1 + E_2 - E}.$$

The $\gamma/J$ relationship will be shown for several sample sizes:

- 10 subjects in each of the two studies
- 16 subjects in study 1, and 32 in study 2 (as in our work)
- 100 subjects in each of the two studies
- 1000 subjects in each of the two studies

For each of the cancer types, the Jaccard index computed from the experimental data is also shown on the graph (as a horizontal reference line). These Jaccard indices are:

- UCEC: $J = 0.036$
- OV: $J = 0.060$
- PRAD: $J = 0.114$

**UCEC**

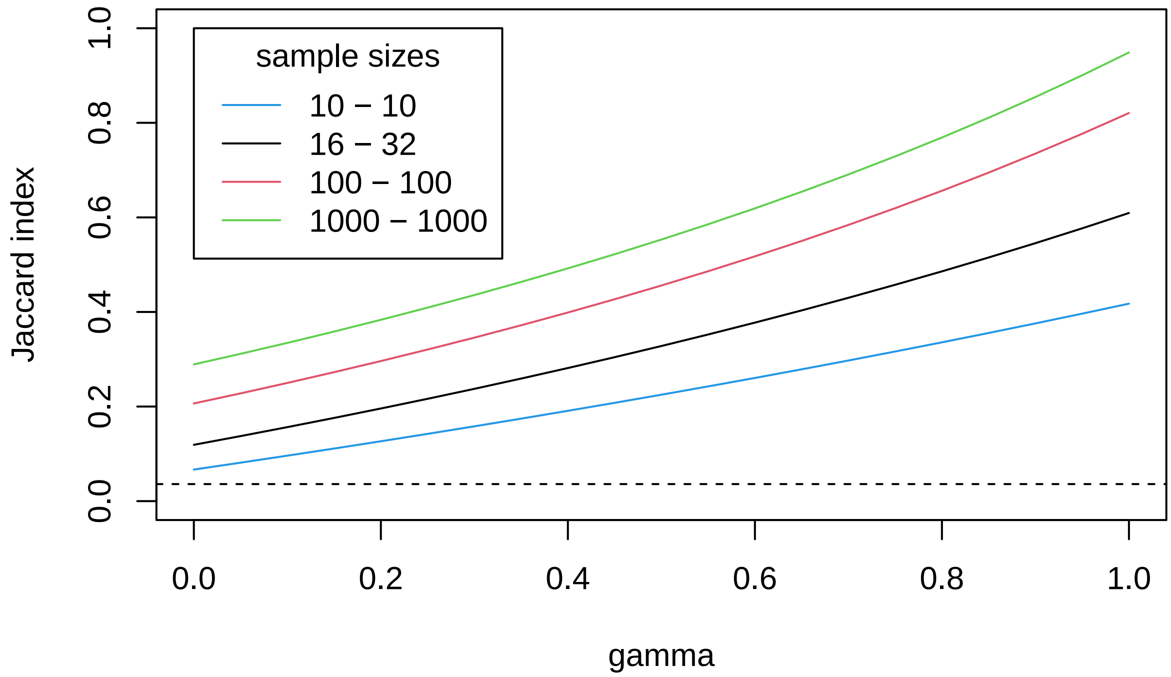
sample sizes
10 − 10
16 − 32
100 − 100
1000 − 1000
Jaccard index
0.0
0.2
0.4
0.6
0.8
1.0
gamma

**OV**

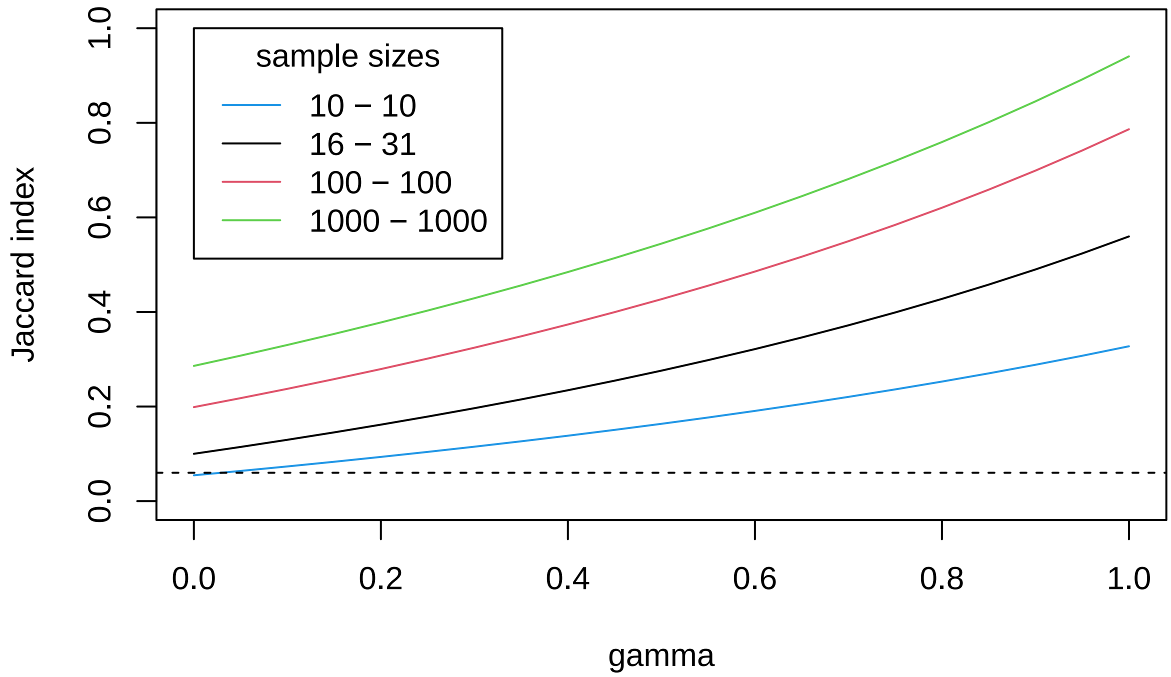
sample sizes
10 − 10
16 − 31
100 − 100
1000 − 1000
Jaccard index
0.0
0.2
0.4
0.6
0.8
1.0
gamma
0.0
0.2
0.4
0.6
0.8
1.0

**PRAD**

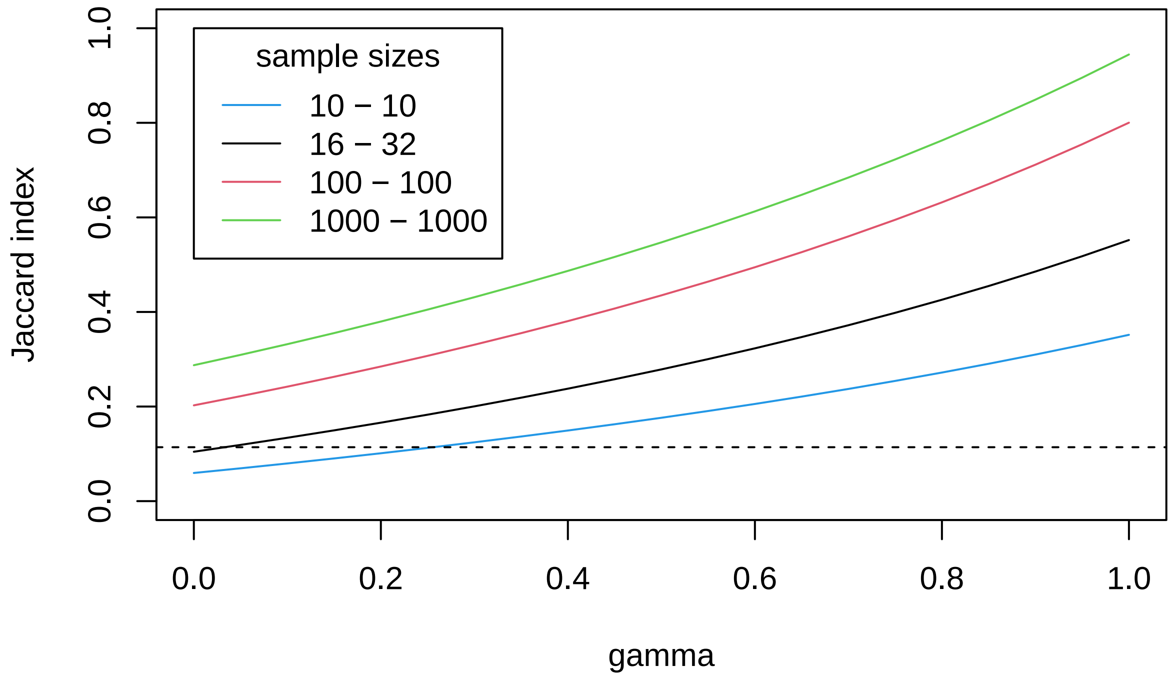


### Discussion

For all three cancer types, the graphs show very similar patterns:

- as the sample size increases the relative overlap between the findings of the two studies increases
- irrespective of the sample size, the relative overlap increases with $\gamma$, i.e. the more similar the two populations, the more DE genes detected by the two methods are expected

In the context of our work, it is the last conclusion that is important: the difference between homogeneous study populations and heterogeneous study populations can even be seen with small sample sizes. The Jaccard indices computed from the experimental data clearly point into the direction of heterogeneous populations.

### Appendix

Let $R_1$ and $R_2$ denote the number of detected DE genes in study 1 and study 2, respectively, and let $R$ number of genes that are detected by both methods. In this appendix expressions for the expected values $E_1 = E(R_1)$, $E_2 = E(R_2)$ and $E = E(R)$ will be developed. Without loss of generality, the calculations are shown for one-sided hypothesis tests.

Upon using the notation introduced earlier, define

$$I_{jk} = 1 \text{ if } H_{0jk} \text{ is rejected, and } I_{jk} = 0 \text{ otherwise.}$$

With this notation, write $(k = 1, 2)$

$$R_k = \sum_{j=1}^{p} I_{jk} \quad \text{and} \quad R = \sum_{j=1}^{p} I_{j1} I_{j2}.$$

The expected value of $R_1$ then becomes

$$\begin{aligned} E(R_1) &= E\left(\sum_{j=1}^{p} I_{j1}\right) = \sum_{j=1}^{p} E(I_{j1}) \\ &= \sum_{j=1}^{p} E\left(E(I_{j1} \mid \theta_1)\right) \\ &= p\, E(\pi(n_1, \sigma_1, \theta_1)), \end{aligned}$$

where $\pi(n_1, \sigma_1, \theta_1)$ is the power of the test for testing $H_{0j1}$. The parameter $\sigma_1$ is the population standard deviation, and $n_1$ is the sample size in the first study. An expression for $E(\pi(n_1, \sigma_1, \theta_1))$ will follow later.

Similarly,

$$\begin{aligned} E(R_2) &= E\left(\sum_{j=1}^{p} I_{j2}\right) = \sum_{j=1}^{p} E(I_{j2}) \\ &= \sum_{j=1}^{p} E\left(E(I_{j2} \mid \theta_2)\right) \\ &= p\, E(\pi(n_2, \sigma_2, \theta_2)). \end{aligned}$$

The power functions, evaluated at a nominal per-comparison significance level of $\alpha$, can be expressed as $(k = 1, 2)$,

$$\pi(n_k, \sigma_k, \theta_k) = P(Z > t_\alpha - \delta_k \mid \theta_k)$$

with $Z$ a standard normal random variable, $t_\alpha$ the critical value of the test at significance level $\alpha$, and $\delta_k$ the non-centrality parameter (ncp), which is here taken to be the ncp of a t-test,

$$\delta_1 = \frac{\sqrt{n_1}}{\sigma_1}\theta_1.$$

With $\theta_1$ distributed as a zero-mean normal random variable with variance $\nu_1$, we find

$$U_1 = Z + \frac{\sqrt{n_1}}{\sigma_1}\theta_1 \sim N\left(0, 1 + \frac{n_1}{\sigma_1^2}\nu_1\right).$$

Hence,

$$\begin{aligned} E(\pi(n_1, \sigma_1, \theta_1)) &= E(P(Z > t_\alpha - \delta_1 \mid \theta_1)) \\ &= P(U_1 > t_\alpha) = 1 - P(U_1 < t_\alpha) \\ &= 1 - H_1(t_\alpha), \end{aligned}$$

with $H_1$ the distribution function of $U_1$. Hence,

$$E(R_1) = p\left[1 - H_1(t_\alpha)\right].$$

For $E(R_2)$ we need the ncp

$$\delta_2 = \frac{\sqrt{n_2}}{\sigma_2}\theta_2 = \frac{\sqrt{n_2}}{\sigma_2}S\theta_1 + \frac{\sqrt{n_2}}{\sigma_2}(1 - S)\theta_2^*.$$

Conditional on $S = 1$, $Z + \delta_2$ is normally distributed with mean zero and variance $1 + \frac{n_2}{\sigma_2^2}\nu_1$, and conditional on $S = 0$, the variance is $1 + \frac{n_2}{\sigma_2^2}\nu_2$, with $\nu_2$ the variance of $\theta_2^*$. Let $H_{21}$ and $H_{20}$ refer to the distribution functions of these two distributions, respectively.

With this notation, and with $P(S = 1) = f$, we find

$$\begin{aligned} E(R_2) &= p\, E(\pi(n_2, \sigma_2, \theta_2)) \\ &= p\, E(P(Z + \delta_2 > t_\alpha)) \\ &= p\, P(Z + \delta_2 > t_\alpha \mid S = 1) f + p\, P(Z + \delta_2 > t_\alpha \mid S = 0)(1 - f) \\ &= pf(1 - H_{21}(t_\alpha)) + p(1 - f)(1 - H_{20}(t_\alpha)). \end{aligned}$$

We now turn to $E(R)$:

$$\begin{aligned} E(R) &= E\left(\sum_{j=1}^{p} I_{j1} I_{j2}\right) \\ &= \sum_{j=1}^{p} E\left(I_{j1} I_{j2}\right) \\ &= pf\, E\left(I_{j1} I_{j2} \mid S = 1\right) + p(1 - f)\, E\left(I_{j1} I_{j2} \mid S = 0\right). \end{aligned}$$

The last expectation becomes (with $Z_1$ and $Z_2$ independent standard normal random variables)

$$\begin{aligned} E\left(I_{j1} I_{j2} \mid S = 0\right) &= P\left(Z_1 > t_\alpha - \frac{\sqrt{n_1}}{\sigma_1}\theta_1 \text{ and } Z_2 > t_\alpha - \frac{\sqrt{n_2}}{\sigma_2}\theta_2^*\right) \\ &= P\left(Z_1 > t_\alpha - \frac{\sqrt{n_1}}{\sigma_1}\theta_1\right) P\left(Z_2 > t_\alpha - \frac{\sqrt{n_2}}{\sigma_2}\theta_2^*\right) \\ &= (1 - H_1(t_\alpha))(1 - H_{20}(t_\alpha)). \end{aligned}$$

The expectation $E\left(I_{j1} I_{j2} \mid S = 1\right)$ can be written as

$$P\left(Z_1 > t_\alpha - \frac{\sqrt{n_1}}{\sigma_1}\theta_1 \ \text{ and } Z_2 > t_\alpha - \frac{\sqrt{n_2}}{\sigma_2}\theta_1\ \right),$$

which is a probability referring to the joint distribution of

$$Z_1 + \frac{\sqrt{n_1}}{\sigma_1}\theta_1 \text{ and } Z_2 + \frac{\sqrt{n_2}}{\sigma_2}\theta_1,$$

which is a bivariate normal distribution with mean zero and variances $1 + \frac{n_1}{\sigma_1^2}\nu_1$ and $1 + \frac{n_2}{\sigma_2^2}\nu_1$ and covariance

$$Cov\left(Z_1 + \frac{\sqrt{n_1}}{\sigma_1}\theta_1, Z_2 + \frac{\sqrt{n_2}}{\sigma_2}\theta_1\right) = \frac{\sqrt{n_1 n_2}}{\sigma_1 \sigma_2}\nu_1.$$

With $H$ the distribution function of this distribution, and upon noting that $H_1$ and $H_{21}$ are the marginal distributions of this bivariate distribution, we find

$$E\left(I_{j1} I_{j2} \mid S = 1\right) = 1 + H(t_\alpha, t_\alpha) - H_1(t_\alpha) - H_{21}(t_\alpha).$$

Hence,

$$E(R) = pf\left[1 + H(t_\alpha, t_\alpha) - H_1(t_\alpha) - H_{21}(t_\alpha)\right] + p(1 - f)(1 - H_1(t_\alpha))(1 - H_{20}(t_\alpha)).$$
