## Supplementary Analysis 2 for "Patient-specific alterations in blood plasma cfRNA profiles enable accurate classification of cancer patients and controls"

**Supplementary Analysis 2 – Biomarker tail genes linked to cancer in literature**

In this study, we identified multiple biomarker tail genes whose abundance in cancer patients' blood plasma or urine strongly deviated from that of cancer-free controls. For the three-cancer cohort, YWHAG, ENO2, MACF1, PIK3CG, and PPIF were some of the tail genes with higher abundance in cancer plasma compared to control plasma. These genes are involved in cancer-related processes such as cell migration, invasion, and metabolism. YWHAG overexpression is linked to worse prognosis and invasiveness in solid tumors and promotes ovarian cancer metastasis[1]. ENO2 **i**s linked to cancer cell proliferation, invasion and metastasis in cancer, including prostate cancer[2,3]. MACF1 plays a crucial role in cell proliferation, migration and cell signaling in various cancers, and high expression is linked to poor prognosis in serous ovarian cancer[4,5]. Higher expression of PPIF, on the other hand, is linked to poor prognosis in uterine cancer[6]. Finally, PIK3CG is implicated in multiple cancers, generally upregulated in cancer tissue and it has been linked to uterine cancer risk and progression[7].

We also detected several genes that were less abundant in the plasma of cancer patients compared to controls in the three-cancer cohort. A few examples of biomarker tail genes with high recurrence among cancer samples are PDCD4, S100A12, RNF126, SF3B6, HIST1H1E, and SNX2. These genes are associated with cancer-related processes, including RNA splicing, inflammation, chromatin structure, and cell signaling.
Note that we sometimes observed opposite directions of RNA abundance in plasma from cancer patients compared to controls, relative to their expression in tissue. PDCD4 is a tumor suppressor known to be downregulated in most solid cancer tissues, aligning with our plasma findings[8]. S100A12 and RNF126, on the other hand, are generally upregulated in cancer tissues and are linked to cancer progression[9–11]. This discrepancy suggests that the mechanisms by which RNAs enter the bloodstream may not be straightforward. Similar findings were reported by Zeka et al.[12], who observed differences in microRNA expression between serum and tissue samples. Further research is needed to fully understand these discrepancies and their implications for cancer diagnostics.

We also detected quite some biomarker tail genes in the lymphoma cohort. Even though not specific for lymphoma, several could be linked to cancer. OTUD5, for example, promotes cancer cell proliferation by deubiquitinating and stabilizing target proteins[13,14]. CTC1 plays an important role in telomere replication and genome stability as part of the CTC1-STN1-TEN1 complex, with decreased CTC1 expression in tumors compared to adjacent normal tissues[15]. MAPKAPK2 (MK2) is involved in the regulation of transcript stability and tumor progression[16]. It is overexpressed in various cancers and is associated with poor prognosis. MYH9 is involved in various cellular processes, including cell adhesion and migration. It can play either an oncogenic or tumor-suppressive role, depending on the type of tumor, tissue environment, and signaling pathways[17].

In the bladder cancer cohort, we only detected 6 consensus biomarker tail genes. For all samples where they were tail genes, the abundance was higher in urine samples from patients compared to controls. CAP1 is known to be involved in actin cytoskeleton remodeling, crucial for cancer cell migration and invasion[18,19]. HSP90AB1 is a molecular chaperone overexpressed in many cancer tissues, including bladder cancer, and is associated with poor prognosis[20]. ALDOA is a glycolytic enzyme linked to cancer cell proliferation and metastasis[21]. TAGLN2 is associated with actin cytoskeleton dynamics and identified as an oncogene[22]. TMSB10 is involved in cell proliferation and migration and overexpressed in many cancer tissues, including bladder cancer[23], and PGK1 promotes tumor development and is linked to chemotherapy resistance and prognosis[24].
