## Supplementary Analysis 3 for "Patient-specific alterations in blood plasma cfRNA profiles enable accurate classification of cancer patients and controls"

**Supplementary Analysis 3 – spikes**

The following plots show spike analyses per cohort. **a** and **b**, Results for sequin spikes added to plasma and urine before RNA isolation. **c** and **d**, Results for ERCC spikes added to RNA eluate after isolation (not added in pan-cancer and three-cancer cohort). **a**, Counts per million (CPM) of Sequin spikes in each sample colored by their abundance in the spike mix (darker, more red colors indicate higher concentration in the mix). Only spikes detected in at least one sample are shown. **b**, Spearman correlations between Sequin spike counts in the samples (DESeq2 normalized). Only spikes with counts above detection threshold in at least half of the samples are shown. **c**, CPM of ERCC spikes colored by their abundance in the spike mix (darker, more red colors indicate higher concentration in the mix). Only spikes detected in at least one sample are shown. **d**, Spearman correlations between ERCC spike counts in the different samples (DESeq2 normalized). Only spikes with counts above detection threshold in at least half of the samples are shown.

We were able to capture the spikes and the high correlation between spikes across samples indicates that they can be used quantitatively. The number of detected spikes varies between cohorts (differences in sequencing depth and/or library preparation method).

<u>Pan-cancer cohort</u>

**a**

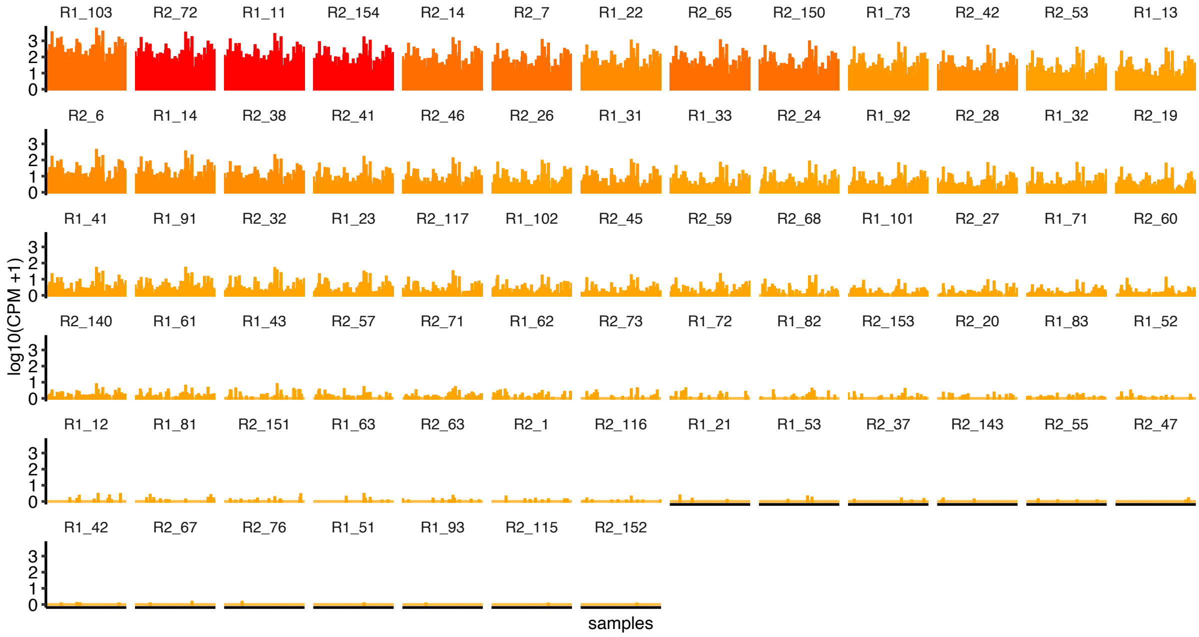


**b**

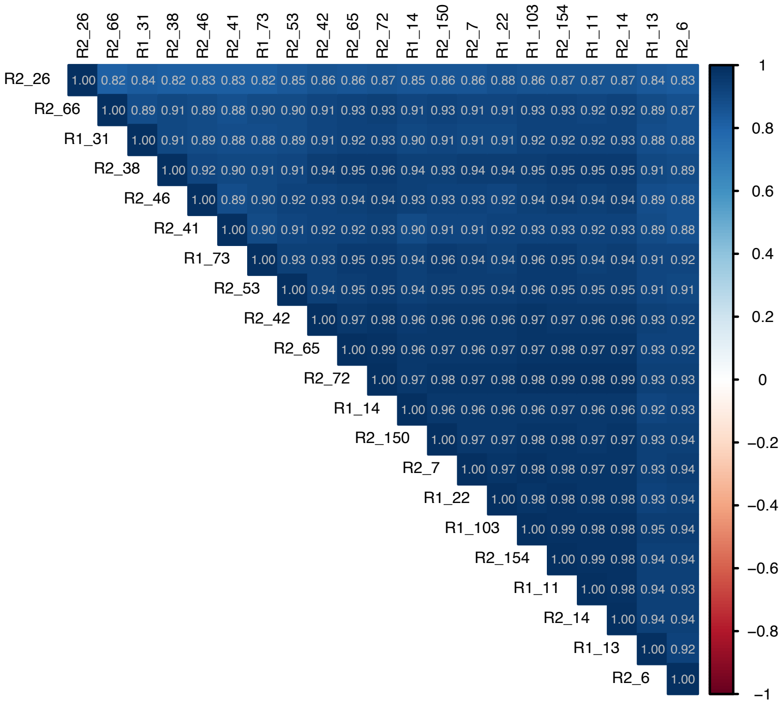

<u>Three-cancer cohort:</u>

**a**

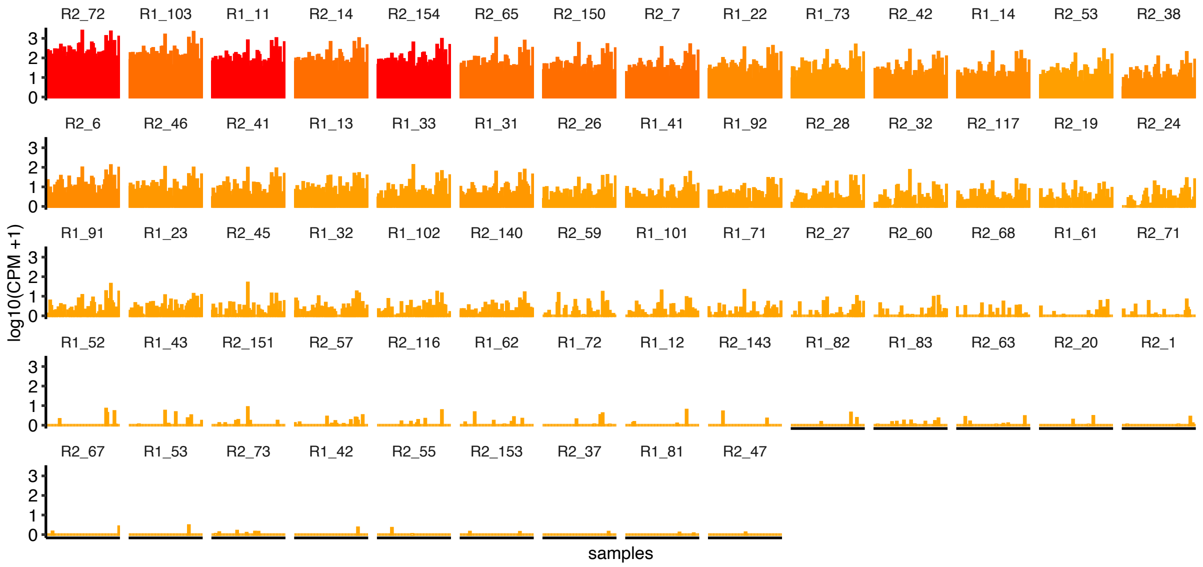


**b**

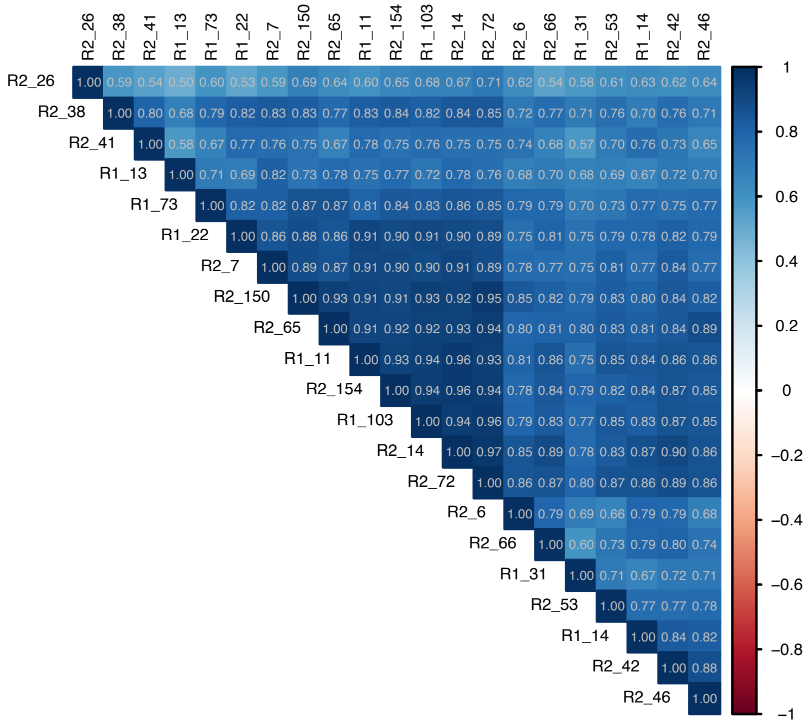

Lymphoma cohort:

**a**

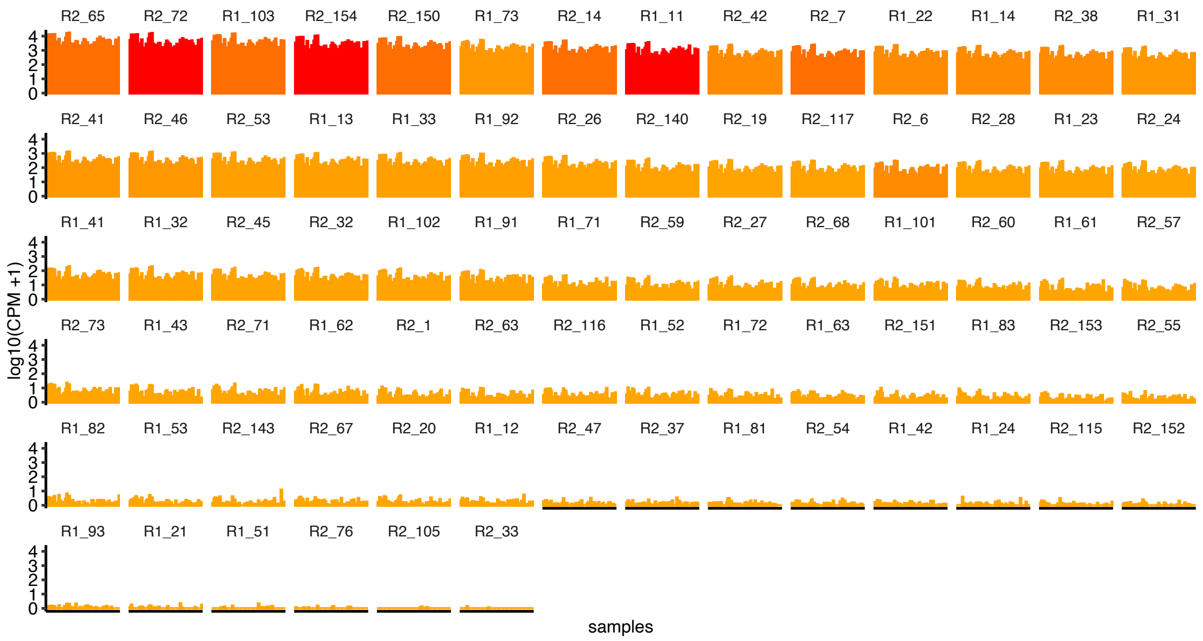


**b**

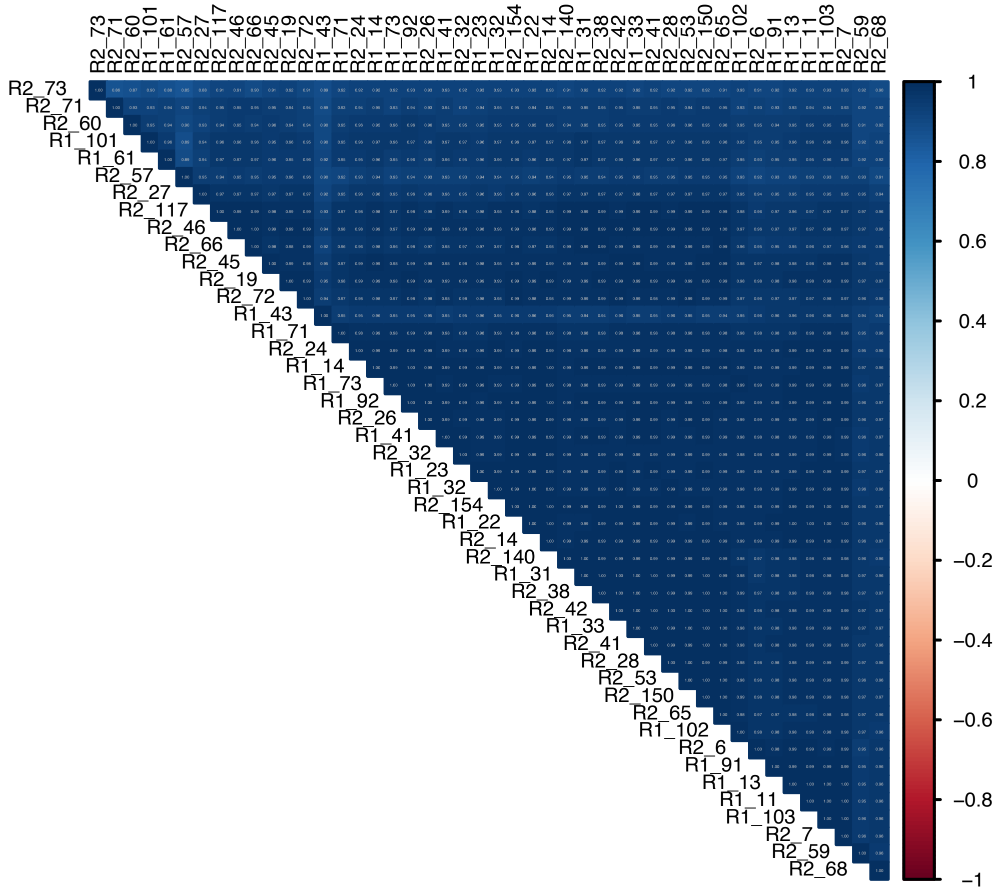

**c**

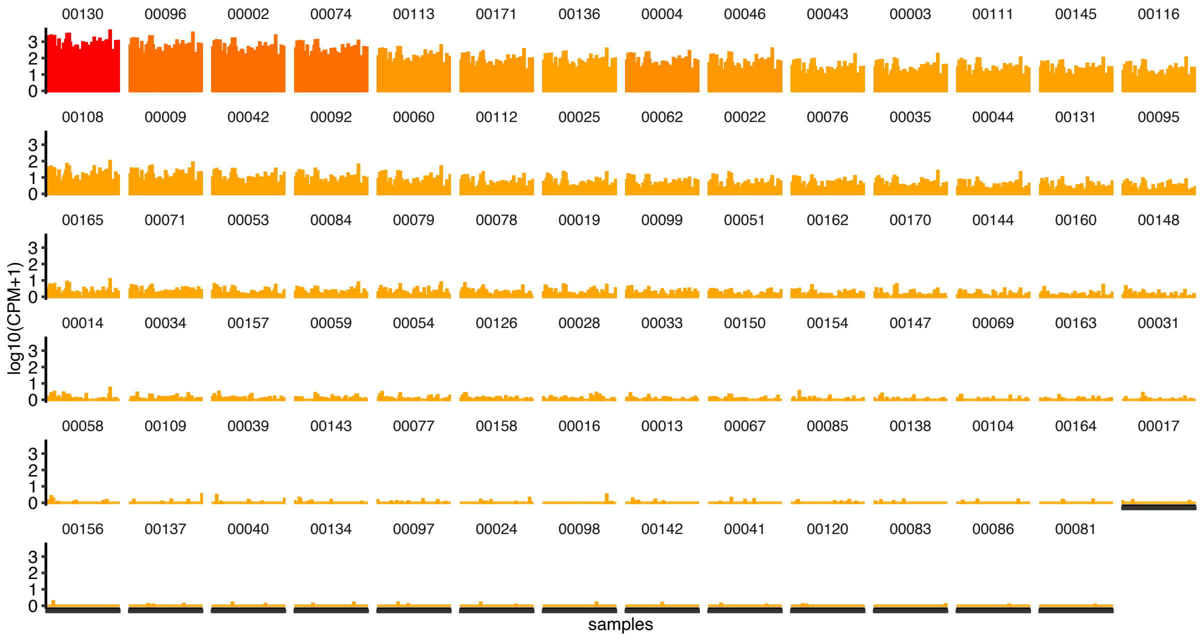
00130 00096 00002 00074 00113 00171 00136 00004 00046 00043 00003 00111 00145 00116
00108 00009 00042 00092 00060 00112 00025 00062 00022 00076 00035 00044 00131 00095
00165 00071 00053 00084 00079 00078 00019 00099 00051 00162 00170 00144 00160 00148
00014 00034 00157 00059 00054 00126 00028 00033 00150 00154 00147 00069 00163 00031
00058 00109 00039 00143 00077 00158 00016 00013 00067 00085 00138 00104 00164 00017
00156 00137 00040 00134 00097 00024 00098 00142 00041 00120 00083 00086 00081
0
1
2
3
log10(CPM+1)
samples

**d**

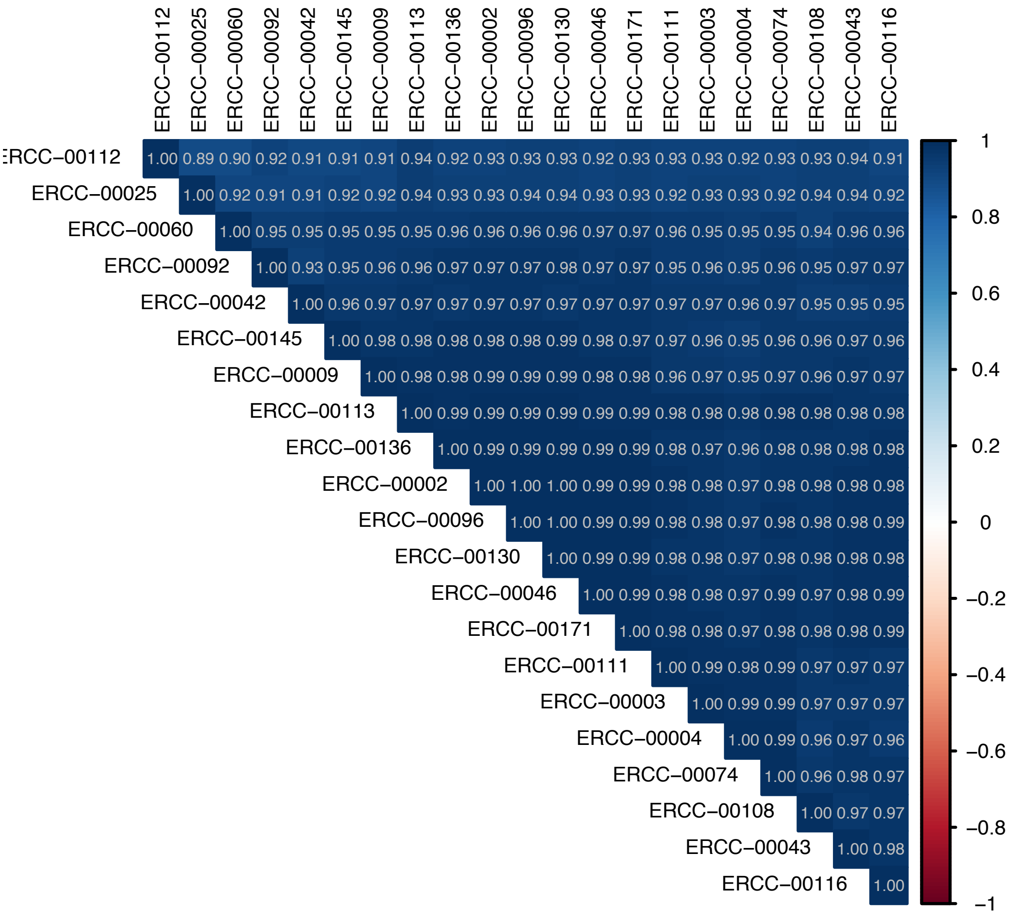
ERCC−00112 ERCC−00025 ERCC−00060 ERCC−00092 ERCC−00042 ERCC−00145 ERCC−00009 ERCC−00113 ERCC−00136 ERCC−00002 ERCC−00096 ERCC−00130 ERCC−00046 ERCC−00171 ERCC−00111 ERCC−00003 ERCC−00004 ERCC−00074 ERCC−00108 ERCC−00043 ERCC−00116
RCC−00112 1.00 0.89 0.90 0.92 0.91 0.91 0.91 0.94 0.92 0.93 0.93 0.93 0.92 0.93 0.93 0.93 0.92 0.93 0.93 0.94 0.91
ERCC−00025 1.00 0.92 0.91 0.91 0.92 0.92 0.94 0.93 0.93 0.94 0.94 0.93 0.93 0.92 0.93 0.93 0.92 0.94 0.94 0.92
ERCC−00060 1.00 0.95 0.95 0.95 0.95 0.95 0.96 0.96 0.96 0.96 0.97 0.97 0.96 0.95 0.95 0.95 0.94 0.96 0.96
ERCC−00092 1.00 0.93 0.95 0.96 0.96 0.97 0.97 0.97 0.98 0.97 0.97 0.95 0.96 0.95 0.96 0.95 0.97 0.97
ERCC−00042 1.00 0.96 0.97 0.97 0.97 0.97 0.97 0.97 0.97 0.97 0.97 0.97 0.96 0.97 0.95 0.95 0.95
ERCC−00145 1.00 0.98 0.98 0.98 0.98 0.98 0.99 0.98 0.97 0.97 0.96 0.95 0.96 0.96 0.97 0.96
ERCC−00009 1.00 0.98 0.98 0.99 0.99 0.99 0.98 0.98 0.96 0.97 0.95 0.97 0.96 0.97 0.97
ERCC−00113 1.00 0.99 0.99 0.99 0.99 0.99 0.99 0.98 0.98 0.98 0.98 0.98 0.98 0.98
ERCC−00136 1.00 0.99 0.99 0.99 0.99 0.99 0.98 0.97 0.96 0.98 0.98 0.98 0.98
ERCC−00002 1.00 1.00 1.00 0.99 0.99 0.98 0.98 0.97 0.98 0.98 0.98 0.98
ERCC−00096 1.00 1.00 0.99 0.99 0.98 0.98 0.97 0.98 0.98 0.98 0.99
ERCC−00130 1.00 0.99 0.99 0.98 0.98 0.97 0.98 0.98 0.98 0.98
ERCC−00046 1.00 0.99 0.98 0.98 0.97 0.99 0.97 0.98 0.99
ERCC−00171 1.00 0.98 0.98 0.97 0.98 0.98 0.98 0.99
ERCC−00111 1.00 0.99 0.98 0.99 0.97 0.97 0.97
ERCC−00003 1.00 0.99 0.99 0.97 0.97 0.97
ERCC−00004 1.00 0.99 0.96 0.97 0.96
ERCC−00074 1.00 0.96 0.98 0.97
ERCC−00108 1.00 0.97 0.97
ERCC−00043 1.00 0.98
ERCC−00116 1.00
1
0.8
0.6
0.4
0.2
0
−0.2
−0.4
−0.6
−0.8
−1

<u>Bladder cancer cohort:</u>

**a**

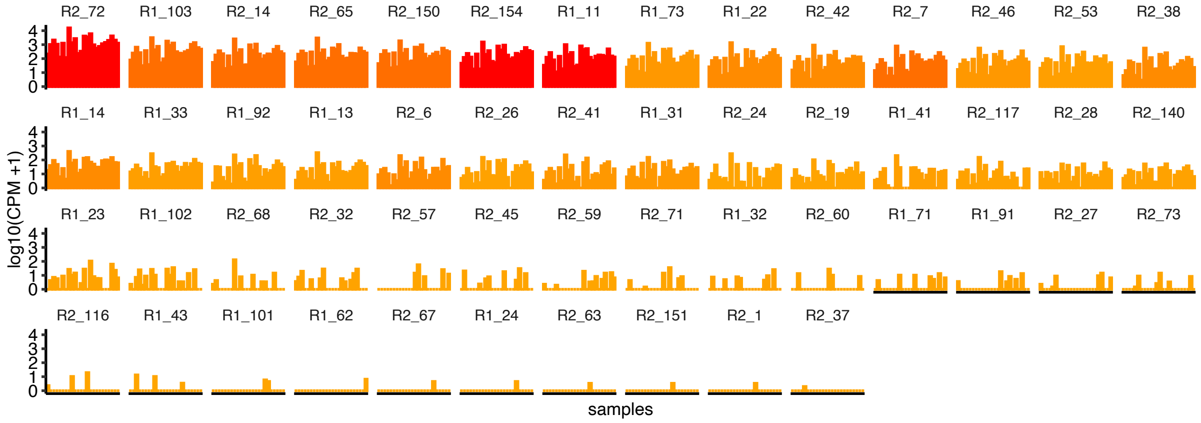


**b**

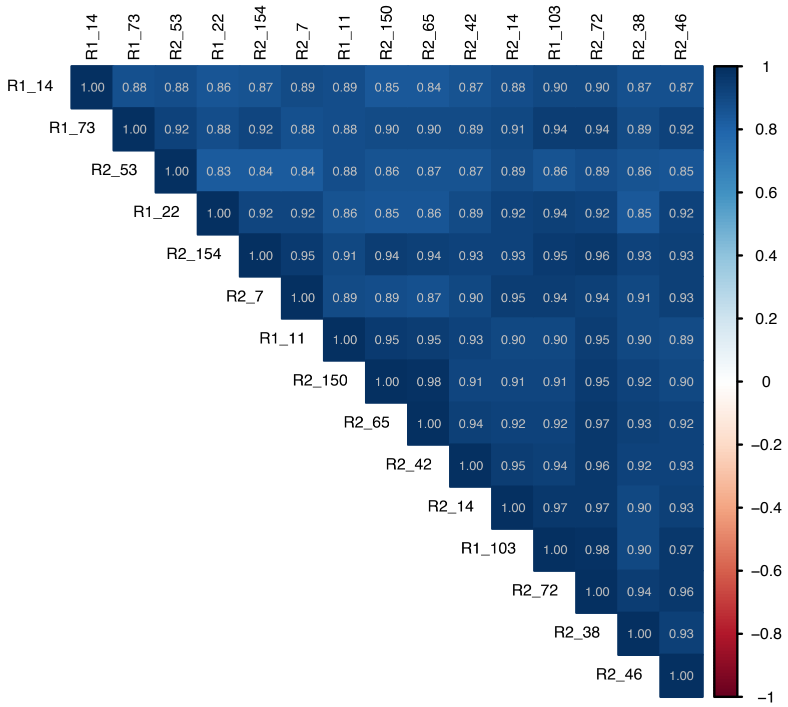

**c**

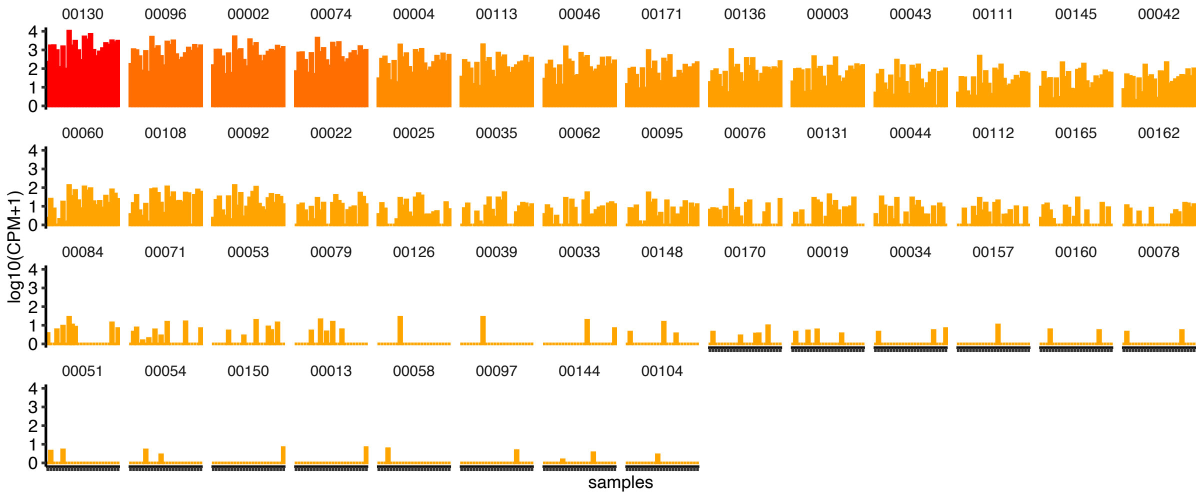
00130 00096 00002 00074 00004 00113 00046 00171 00136 00003 00043 00111 00145 00042
00060 00108 00092 00022 00025 00035 00062 00095 00076 00131 00044 00112 00165 00162
00084 00071 00053 00079 00126 00039 00033 00148 00170 00019 00034 00157 00160 00078
00051 00054 00150 00013 00058 00097 00144 00104
0 1 2 3 4
log10(CPM+1)
samples

**d**

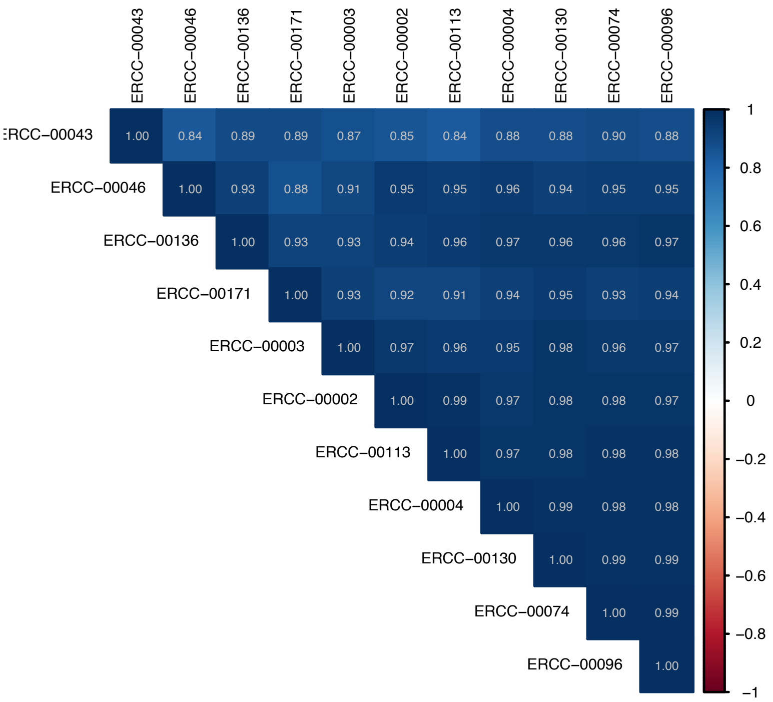
ERCC−00043 ERCC−00046 ERCC−00136 ERCC−00171 ERCC−00003 ERCC−00002 ERCC−00113 ERCC−00004 ERCC−00130 ERCC−00074 ERCC−00096
ERCC−00043 1.00 0.84 0.89 0.89 0.87 0.85 0.84 0.88 0.88 0.90 0.88
ERCC−00046 1.00 0.93 0.88 0.91 0.95 0.95 0.96 0.94 0.95 0.95
ERCC−00136 1.00 0.93 0.93 0.94 0.96 0.97 0.96 0.96 0.97
ERCC−00171 1.00 0.93 0.92 0.91 0.94 0.95 0.93 0.94
ERCC−00003 1.00 0.97 0.96 0.95 0.98 0.96 0.97
ERCC−00002 1.00 0.99 0.97 0.98 0.98 0.97
ERCC−00113 1.00 0.97 0.98 0.98 0.98
ERCC−00004 1.00 0.99 0.98 0.98
ERCC−00130 1.00 0.99 0.99
ERCC−00074 1.00 0.99
ERCC−00096 1.00
1 0.8 0.6 0.4 0.2 0 −0.2 −0.4 −0.6 −0.8 −1
